# Inequalities in early childhood developmental concerns before, during and after the COVID-19 pandemic in Scotland: a retrospective cohort study

**DOI:** 10.64898/2026.04.30.26352025

**Authors:** Iain Hardie, Louise Marryat, Aja Murray, Josiah King, Kenneth Okelo, Lynda Fenton, James P. Boardman, Michael V. Lombardo, Philip Wilson, Rachael Wood, Bonnie Auyeung

## Abstract

**Background:** The COVID-19 pandemic was associated with increased child developmental concerns in Scotland. However, it is not known whether this increase was uniform across social groups, and there is particular concern that children from low-income households, urban areas and ethnic minority groups may have been disproportionately affected. This retrospective, population-based, cohort study aimed to examine whether the pandemic was associated with changes in developmental inequalities in Scotland.

**Study design:** We linked national birth records, the COVID-19 in Pregnancy in Scotland (COPS) dataset, and 13-15 month and 27-30 month child health review records, covering all children born in Scotland who undertook reviews between January 2019 and August 2023 and had full developmental data. Logistic regression models estimated inequalities in odds of developmental concerns, before, during and after the pandemic and across Scottish Index of Multiple Deprivation (SIMD) quintiles, parental National Statistics Socioeconomic Classification (NS-SEC), urban-rural classification, child ethnicity and child sex. Interaction analysis formally tested for any significant changes in inequalities.

**Findings:** The analyses included 254,367 children, covering 13-15 month child health review records for 183,439 children and 27-30 month child health review records for 184,689 children. Children in more deprived SIMD quintiles and lower parental NS-SEC categories had significantly higher odds of developmental concerns, as did African and Asian children (at 27-30 months only). Children who were female and in rural areas (27-30 months only) had significantly lower odds of developmental concerns. Developmental inequalities were broadly consistent at each time point and interaction analysis suggested that there was no widening of inequalities during or after the pandemic.

**Conclusions:** Developmental inequalities in Scotland did not widen during or after the COVID-19 pandemic. However, substantial pre-existing inequalities persist, underscoring the need for interventions to reduce disparities and support national policy goals on child development.

## Introduction

Early childhood is a critical period for cognitive, social, emotional, behavioural and motor development. It represents a sensitive time for brain development during which environmental and social factors, along with biological factors, act together to shape long-term developmental outcomes [1]. In Scotland, as in many other countries, developmental outcomes vary between children from different sociodemographic backgrounds. One key aspect of this is socioeconomic inequalities, with children from lower socioeconomic groups generally experiencing poorer outcomes [2]. This arises through multiple interacting pathways, including family level factors such as parental mental health, parental education, adverse childhood experiences, family functioning and attachment, and wider economic, social, environmental and multi-level factors such as household income, deprivation, childcare quality, peer relationships, access to services, nutrition, pollution, climate change and discrimination [1-4]. Beyond socioeconomic gradients, UK data also indicate developmental inequalities by sex and ethnicity. For example, early cognitive developmental outcomes tend to be poorer among boys than girls, and poorer among Black, Pakistani and Bangladeshi children than among White children [5]. Evidence on urban-rural differences is sparser. Some limited data suggest fewer emotional development problems among children from rural areas in England, but no urban-rural differences in Scotland [6, 7].

During the COVID-19 pandemic, countries around the world implemented stringent Public Health and Social Measures (PHSM) to reduce viral transmission. In Scotland, these included ‘stay at home’ orders, strict restrictions on households, closure of recreational facilities, limits on outdoor hours and, for most children, closure of nurseries, schools, early learning and childcare settings [8]. Although effective in limiting infection spread [9], these measures may have had unintended developmental consequences. PHSM reduced peer interaction important for language, cognitive, and socioemotional development [10], restricted access to centre-based childcare, which confers cognitive benefits particularly for disadvantaged children [11], and placed financial and psychological strain on families, potentially affecting parents’ capacity to provide nurturing care [1]. Conversely, exposure to PHSM may in some ways have improved childhood development, for example by fostering closer parent-child relationships.

Overall, emerging evidence suggests that the pandemic was generally associated with poorer early developmental outcomes, with effects appearing more closely linked to PHSM than to SARS-CoV-2 infection itself. For example, in Scotland, analysis of linked administrative health data indicates that PHSM were associated with increased developmental concerns at a population level, whereas prenatal SARS-CoV-2 exposure was not significantly associated with early developmental outcomes at an individual level [12, 13]. Adverse pandemic PHSM impacts have also been observed internationally [14].

Importantly, these impacts may not have been evenly distributed across society. Children from low-income households may have been particularly vulnerable. This is because withdrawal of centre-based childcare under PHSM may have disproportionately affected disadvantaged children, who are often those who benefit most from high-quality provision [11]. Moreover, the pandemic may have increased financial insecurity faced by parents, for example if they faced income losses due to PHSM. This may have impacted children due to known links between parents’ wellbeing and child development [15]. Urban children may also have experienced greater disruption, as restrictions were often stricter and longer-lasting in Scottish cities (particularly Glasgow) due to higher infection rates [8], whereas lower population density in rural areas may have mitigated some social disruption. There were also fewer geographical constraints on movement in urban areas, and likely fewer families impacted by business closures.

International evidence on whether the pandemic widened developmental inequalities is mixed. One review suggests exacerbation of pre-existing racial and socioeconomic disparities in children’s behavioural health [16], and several quantitative studies from Canada, the US, Germany, India, South Korea, Bangladesh and Uruguay report stronger negative impacts among socioeconomically disadvantaged children [17-24]. Conversely, the findings of other quantitative studies from the UK and Germany suggest that socioeconomic inequalities in child development and mental health may have actually decreased during the pandemic due to disadvantaged groups experiencing less pronounced impacts than other groups [25, 26]. There is currently a lack of large, population-based studies. Moreover, the impacts of the pandemic are likely to have varied between countries with differing approaches to PHSM and with differing health, education and social systems. As such, it is important that research is carried out in a wide-range of countries in order to compare evidence internationally.

The present study, conducted as part of the wider COVID-19 Health Impact on Long-term Child Development in Scotland (CHILDS) project, aimed to assess developmental inequalities in Scotland, before, during and after the COVID-19 pandemic using a population-based retrospective cohort design.

## Methods

### Study design and participants

We conducted a population-based retrospective cohort study, comparing odds of developmental concerns among all children aged 13-15 months or 27-30 months in Scotland before, during and after the COVID-19 pandemic and across socioeconomic measures, urban-rural classification, child ethnicity and child sex. This was carried out by utilising Scottish administrative health data, linking the COVID-19 in Pregnancy in Scotland (COPS) dataset, child birth records collected by National Records for Scotland (NRS), routine 13-15 month and 27-30 month child health review records and maternal Mental Health Inpatient and Day Case Scottish Morbidity Records (SMR04) [27-30].

Child health reviews are universal assessments delivered by trained health visitors, who provide specialist advice and support to families (e.g. often acting as a gateway to referral for additional support). Coverage of these reviews is consistently high (including during the pandemic) and does not vary across key characteristics like child sex and socioeconomic deprivation [31]. Our study covered all children who completed a 13–15 month or 27–30 month health review between January 2019 and August 2023 and had complete data on developmental outcomes and key socioeconomic variables. In total, 200,424 children had a recorded 13-15 month review and 208,028 children had a recorded 27-30 month review. Of these, we excluded 16,985 13-15 month review records and 23,339 27-30 month review records due to incomplete developmental assessments or missing key sociodemographic data. Therefore, our final analysis included 183,439 13-15 month reviews and 184,689 27-30 month reviews, representing 254,367 individual children in total. This represents approximately 80% of all children in Scotland who were eligible for these reviews during our analysis period.

### Procedures

#### Developmental Outcomes

Two binary outcome variables were derived: (1) any (i.e. at least one) child developmental concerns identified at 13-15 month child health reviews, and (2) any (i.e. at least one) child developmental concerns identified at 27-30 month child health reviews. Outcomes were coded as presence or absence of any developmental concerns (including newly or previously identified concerns). Concerns could relate to speech-language-communication, problem solving, gross motor, fine motor, personal-social, or emotional-behavioural development. Health visitors identified concerns based on discussions with parents/caregivers, structured observation of the child, and the results of Ages and Stages Questionnaires 3rd Edition (ASQ-3).

#### Pandemic cohort classification

Children were categorised according to the timing of their health review relative to the pandemic, as follows: (1) pre-pandemic cohort, consisting of reviews between 1^st^ January 2019 and 22^nd^ March 2020, (2) pandemic cohort, consisting of reviews between 23^rd^ March 2020 and 9^th^ August 2021, and (3) post-pandemic cohort, consisting of reviews between 10^th^ August 2021 and 31^st^ August 2023. These reflect the periods immediately before, during and after key PHSM were in place in Scotland [8].

#### Inequality measures

Developmental inequalities were examined across five inequality measures. Firstly, Scottish Index of Multiple Deprivation (SIMD) was used as a measure of area-based deprivation. This reflects aggregate information on income, employment, education, health, access to services, crime, and housing in small administrative areas across Scotland. Children were assigned to SIMD quintiles based on their postcode of residence as measured at child health reviews (i.e. quintile 1=most deprived to 5=least deprived). Secondly, parental National Statistics Socioeconomic Classification (NS-SEC) was used as a socioeconomic status measure. This was derived from parental occupation recorded at time of birth in NRS birth records. It was grouped into four categories: not employed, routine/manual, intermediate, and managerial/professional, and was based on the highest available NS-SEC classification of parental occupation. Thirdly, the Scottish Government’s two-fold Urban Rural Classification was used to distinguish between children in urban and rural areas [32]. This was based on their home postcode as recorded in the COPS dataset. Fourthly, child ethnic group was used to assess inequalities across ethnicity. This was recorded at health reviews and categorised as White, Mixed, Asian, African, Caribbean/Black, or Other. Finally, child sex (male/female) as recorded in birth records was used to assess sex differences.

#### Control variables

Our primary analysis also included three confounders and covariates as control variables. These were as follows: (1) maternal age at childbirth (continuous, in years) as measured in maternity hospital records, (2) maternal mental health hospital admissions, which came from maternal hospital records and was used as a proxy for severe mental ill-health, and (3) maternal smoking status as recorded in antenatal booking records. These were selected as they are key factors influencing child development. Finally, an additional sensitivity analysis controlled for gestational age at birth (continuous, in weeks).

### Statistical analysis

We first examined the distribution of key variables by pandemic period and child age at assessment using descriptive statistics. We then plotted the proportion of children with developmental concerns over time (pre-pandemic, during the pandemic, and post-pandemic), stratified by SIMD quintile, parental NS-SEC, urban–rural classification, child ethnic group, and child sex. Logistic regression models were used to estimate inequalities in the odds of developmental concerns across each inequality measure. Analyses were conducted separately for the pre-pandemic cohort, pandemic, and post-pandemic cohorts. Coefficient plots were generated to visualise differences in the magnitude of inequalities across periods. This modelling was conducted in two stages. Firstly, unadjusted models examined each inequality measure separately (i.e. in separate bivariate models). Secondly, adjusted models included SIMD quintile, parental NS-SEC, urban-rural classification, child ethnic group and child sex simultaneously, with additional adjustment for maternal age, maternal mental health hospital admissions and maternal smoking status as control variables. In addition, to assess more formally whether the pandemic was associated with any significant changes in developmental inequalities, an additional step of analysis examined interaction between pandemic period (pre-, during, post-) and each of the inequality measures. These interaction models were also estimated in both unadjusted and fully adjusted forms.

Finally, in addition to the main analysis, two sensitivity analyses were also conducted. Firstly, the primary analyses were repeated with children from the Greater Glasgow and Clyde health board excluded. This was to ensure known data issues in this health board (lower 13-15 month health review uptake prior to August 2019 and incomplete 27-30 month problem-solving developmental data prior to May 2019) did not impact our findings. Secondly, the primary analyses models were re-estimated with adjustment for gestational age at birth. This was to assess the direct relationship between pandemic period and inequalities not mediated through any association between pandemic period and gestational age.

All analyses were conducted within Scotland’s National Safe Haven using Stata IC version 16.1. Our analysis plan was preregistered using Open Science Framework (https://osf.io/awfev). All results are reported in accordance with the RECORD guidelines (see supplementary appendix Table A1 for RECORD checklist).

## Results

Descriptive statistics, stratified by pandemic period and age at assessment, are provided in Table 1. Overall, 19,719 (10.7%) children had developmental concerns identified at 13-15 month reviews, whilst 27,741 (15.0%) had developmental concerns identified at 27-30 month reviews. The higher prevalence at 27-30 months reflects greater detectability of developmental difficulties as children age. Among children assessed at 13-15 months, 93,978 (51.2%) were male and 89,461 (48.8%) were female, whilst 165,435 (90.2%) were recorded as belonging to a white ethnic group and 18,004 (9.8%) from other ethnic groups. Among those assessed at 27-30 months, 94,106 (51.0%) were male and 90,583 (49.0%) were female, whilst 168,050 (91.0%) children were recorded as white and 16,639 (9.0%) children from other ethnic groups.

**Table 1.**
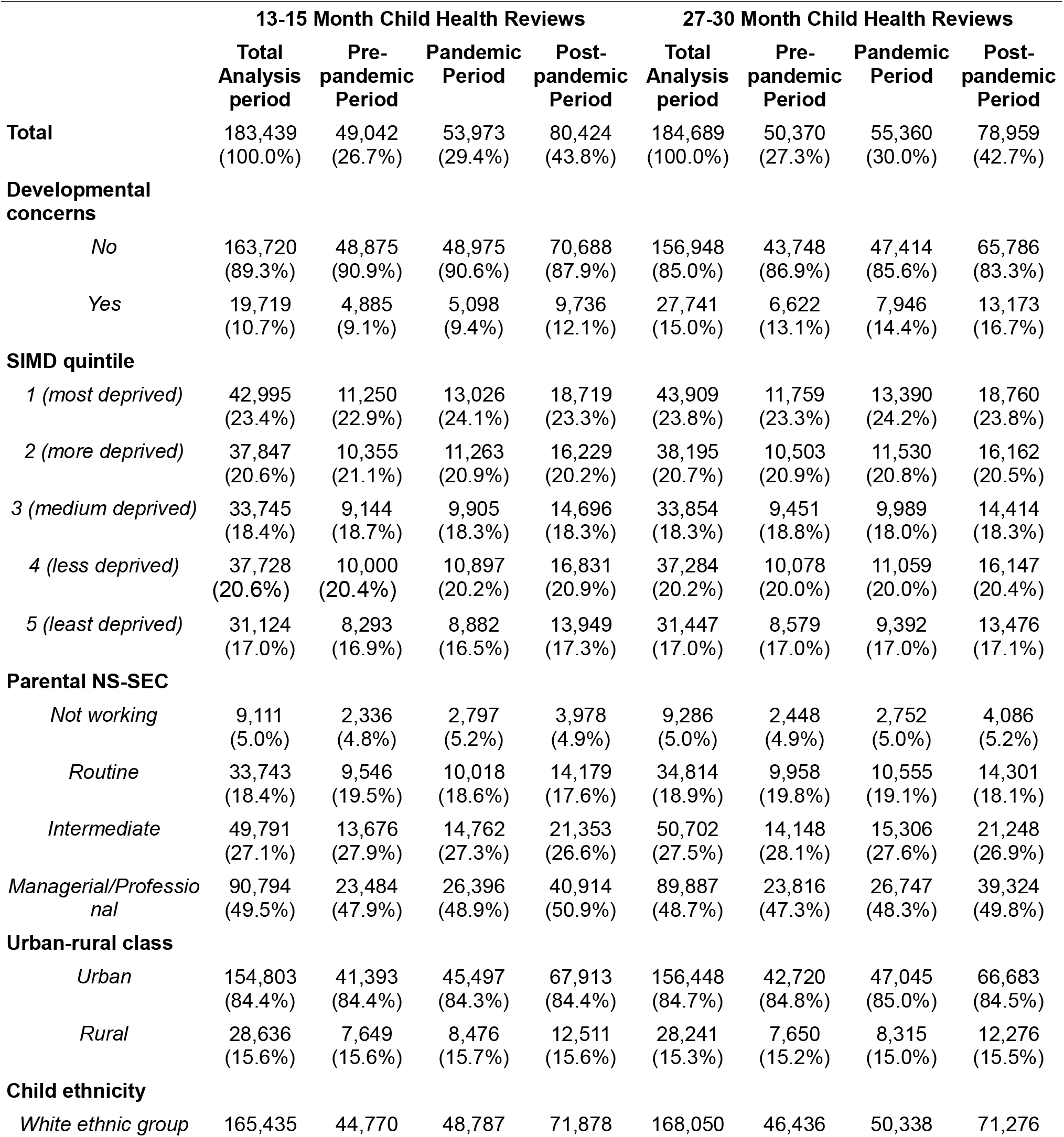
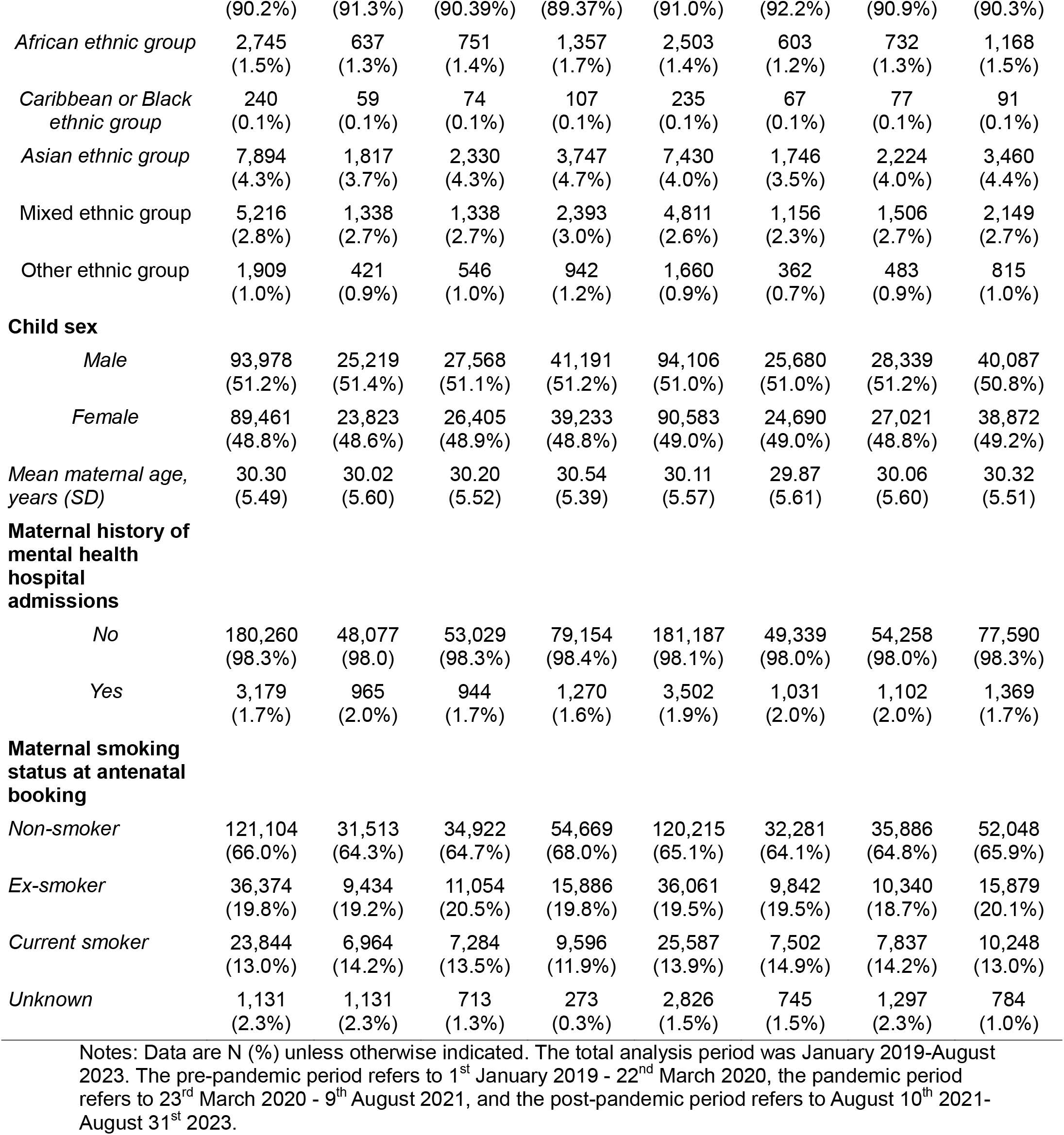
Descriptive statistics on all analysis variables for study children by age of health review assessment and cohort.

Descriptive plots illustrating the proportion of children with developmental concerns over time, stratified by SIMD quintile, parental NS-SEC, urban-rural classification, child ethnic group and child sex are provided in Figures 1 and 2. Overall, the figures demonstrate clear developmental inequalities. Higher proportions of concerns were observed among male children, those from more deprived areas, and those whose parents were not working or were in routine/manual occupations. Inequalities were more pronounced at 27-30 month reviews than 13-15 month reviews. Urban-rural differences were not evident at 13-15 months, however at 27-30 months children living in urban areas showed a higher proportion of concerns. Patterns by ethnicity varied by age. At age 13-15 months, White children had a slightly higher proportion of concerns, whereas at 27-30 months children from other ethnic backgrounds showed higher proportions. In line with previous research [13], the overall proportion of children with developmental concerns increased following the introduction of COVID-19 PHSM before stabilising (but remaining higher than pre-pandemic levels) after restrictions were lifted. Importantly, this increase appeared broadly consistent across social groups.

**Figure 1.**
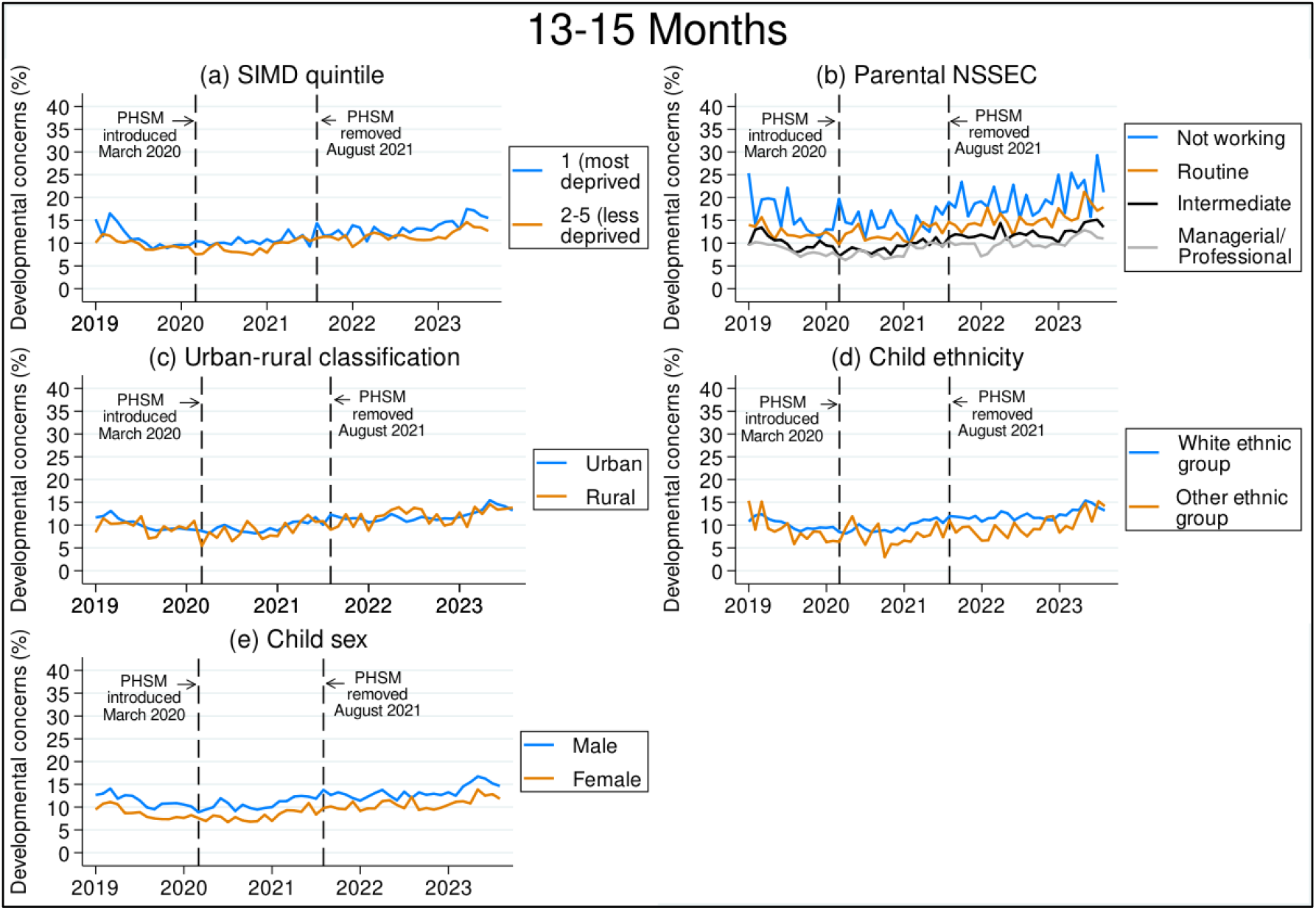
Proportion of children with developmental concerns at age 13-15 months by SIMD quintile, parental NS-SEC, urban-rural classification, child ethnic group and child sex, January 2019-August 2023.

**Figure 2.**
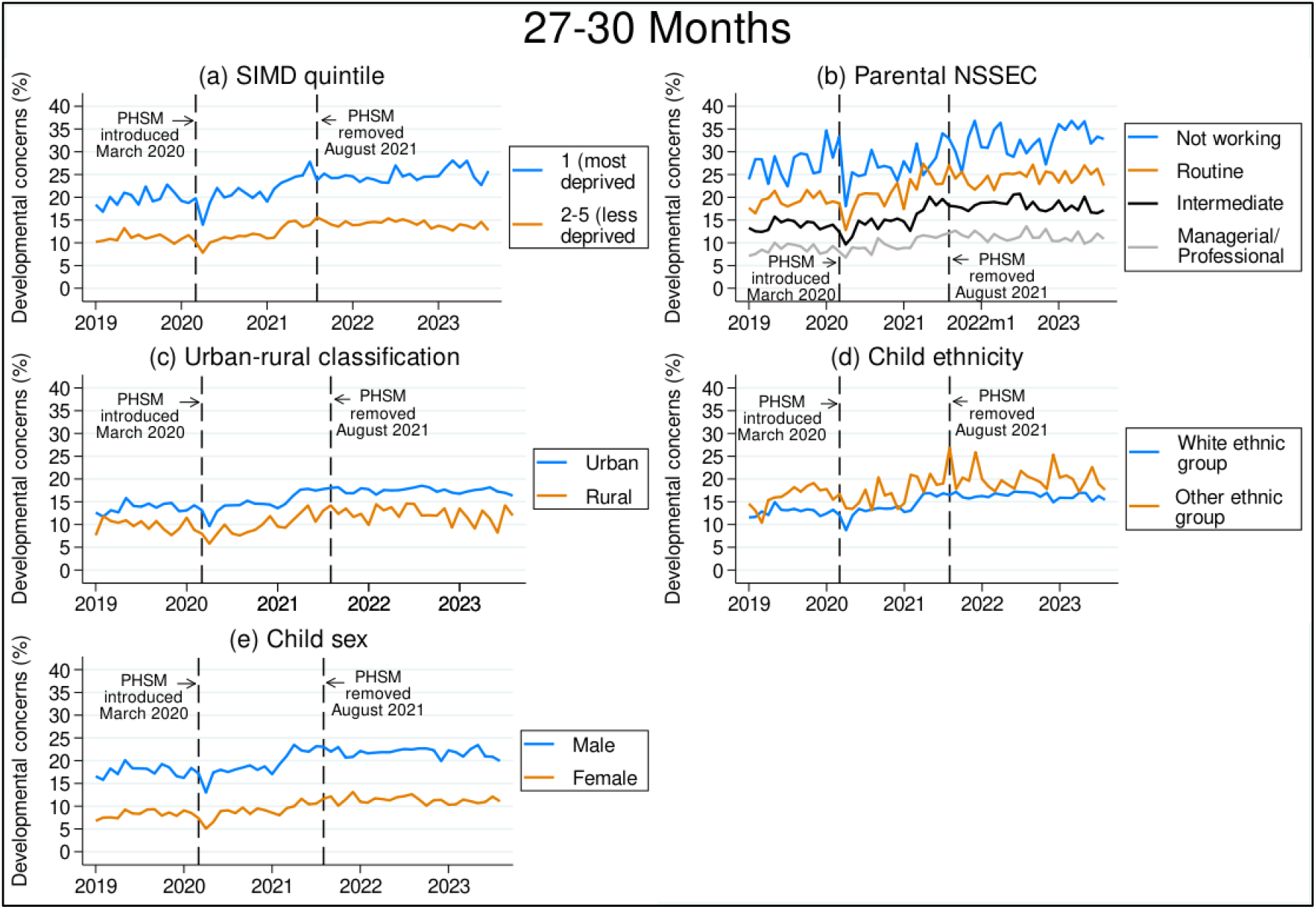
Proportion of children with developmental concerns at age 27-30 months by SIMD quintile, parental NS-SEC, urban-rural classification, child ethnic group and child sex, January 2019-August 2023.

Results from the adjusted logistic regression models, estimated separately for the pre-pandemic, pandemic, and post-pandemic cohorts, are provided in Figures 3 and 4. In addition, results of the unadjusted models are provided in supplementary appendix Figures A1 and A2, and the results are also provided in table-form in supplementary appendix Table A2. As with the descriptive analysis (Figures 1-2), the results highlight clear developmental inequalities across SIMD, parental NS-SEC and child sex, with (1) significantly higher odds of developmental concerns among children from more deprived areas (compared to the least deprived reference category), (2) significantly higher odds of developmental concerns among children whose parents were not working or in routine or intermediate jobs (compared to the managerial/professional reference category), and (3) significantly lower odds of developmental concerns among female children (compared to the male reference category). In addition, children in rural areas had significantly lower odds of developmental concerns (compared to urban reference category) at 27-30 months but not 13-15 months. Associations with ethnicity were mixed at age 13-15 months, but African and Asian children had consistently and significantly higher odds of developmental concerns at age 27-30 months. Overall, the magnitude and pattern of developmental inequalities appeared to be broadly consistent over time when comparing inequalities between the pre-pandemic, pandemic and post-pandemic cohorts.

**Figure 3.**
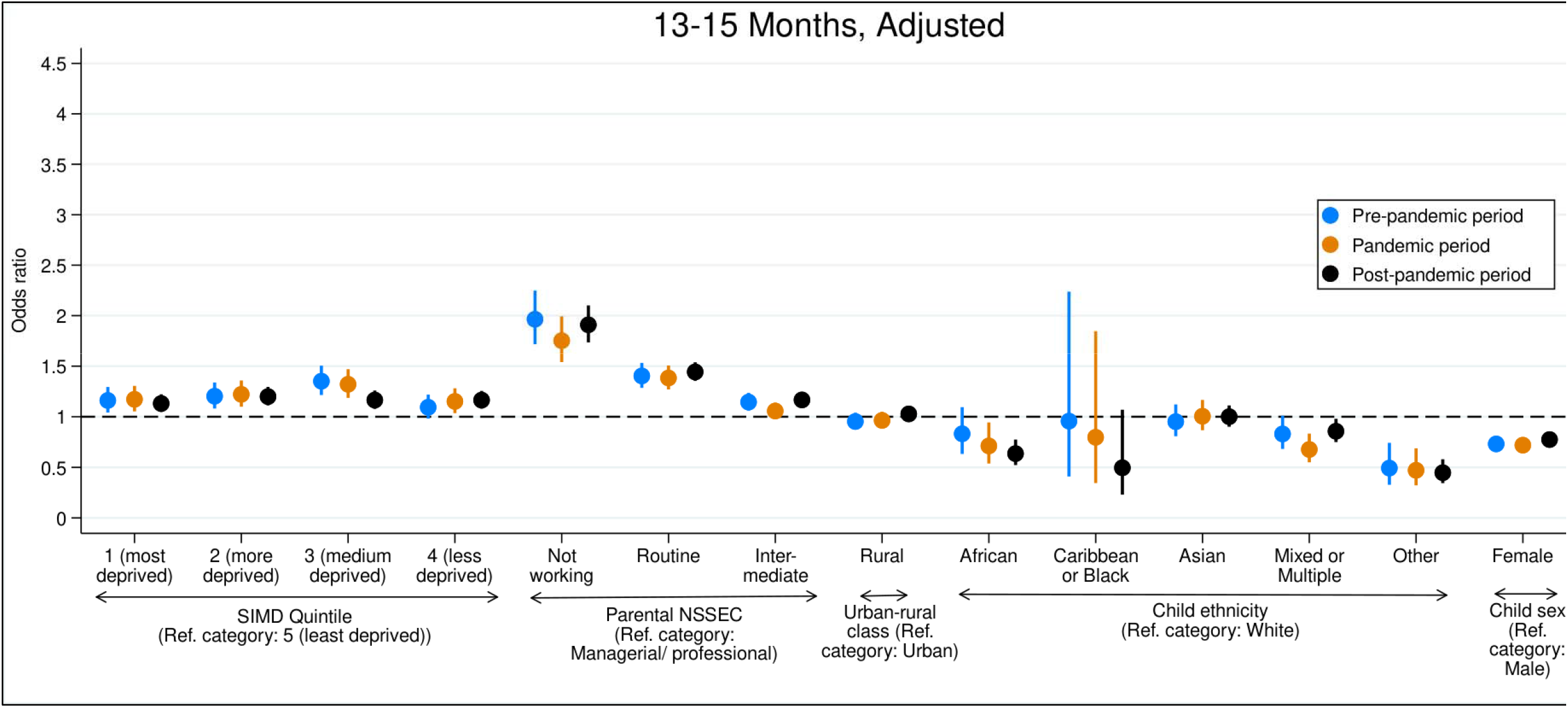
Adjusted odds ratios and 95% confidence intervals showing inequalities in odds of developmental concerns at age 13-15 months before, during and after the COVID-19 pandemic. Notes: odds ratios are shown by point estimates and 95% confidence intervals are shown by vertical bars. The dotted line indicates no difference in odds ratios compared to reference groups, whilst higher odds ratios above this indicate increased odds of developmental concerns compared to reference groups. Models were conducted separately for the pre-pandemic cohort, pandemic cohort and post-pandemic cohort, and odds ratios can be compared to see how inequalities differed over time between cohorts. Models adjust for other inequality measures as well as maternal age, maternal mental health hospital admissions and maternal smoking status as additional control variables.

**Figure 4.**
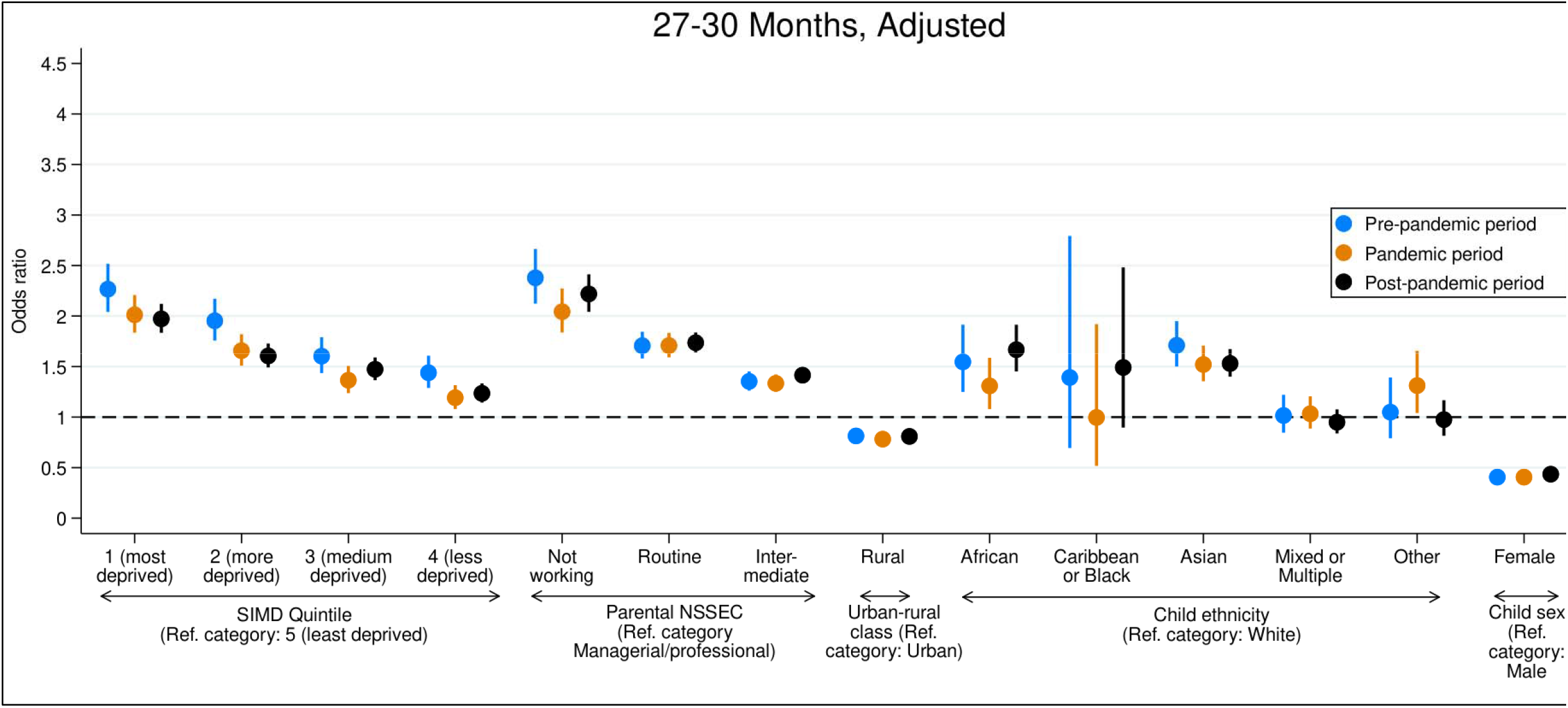
Adjusted odds ratios and 95% confidence intervals showing inequalities in odds of developmental concerns at age 27-30 months before, during and after the COVID-19 pandemic. Notes: odds ratios are shown by point estimates and 95% confidence intervals are shown by vertical bars. The dotted line indicates no difference in odds ratios compared to reference groups, whilst higher odds ratios above this indicate increased odds of developmental concerns compared to reference groups. Models were conducted separately for the pre-pandemic cohort, pandemic cohort and post-pandemic cohort, and odds ratios can be compared to see how inequalities differed over time between cohorts. Models adjust for other inequality measures as well as maternal age, maternal mental health hospital admissions and maternal smoking status as additional control variables.

Results of the interaction analysis, which more formally assessed whether the pandemic was associated with any significant changes in developmental inequalities, are provided in Table 2. At 13-15 months, there was no significant interaction between the pandemic cohort variable and any of the inequality measures. This indicates that developmental inequalities across SIMD, parental NS-SEC, urban-rural classification child ethnicity or child sex did not significantly widen or narrow during the pandemic or post-pandemic periods when compared to the pre-pandemic period. Similarly, at age 27-30 months there was no significant interaction between the pandemic cohort variable and most inequality measures, again suggesting no substantial change in the magnitude of developmental inequalities over time. The only exception was SIMD. Inequalities between children in the least deprived quintile and those in the middle or more deprived quintiles showed some narrowing during the pandemic and post-pandemic periods when compared to the pre-pandemic period.

**Table 2.**
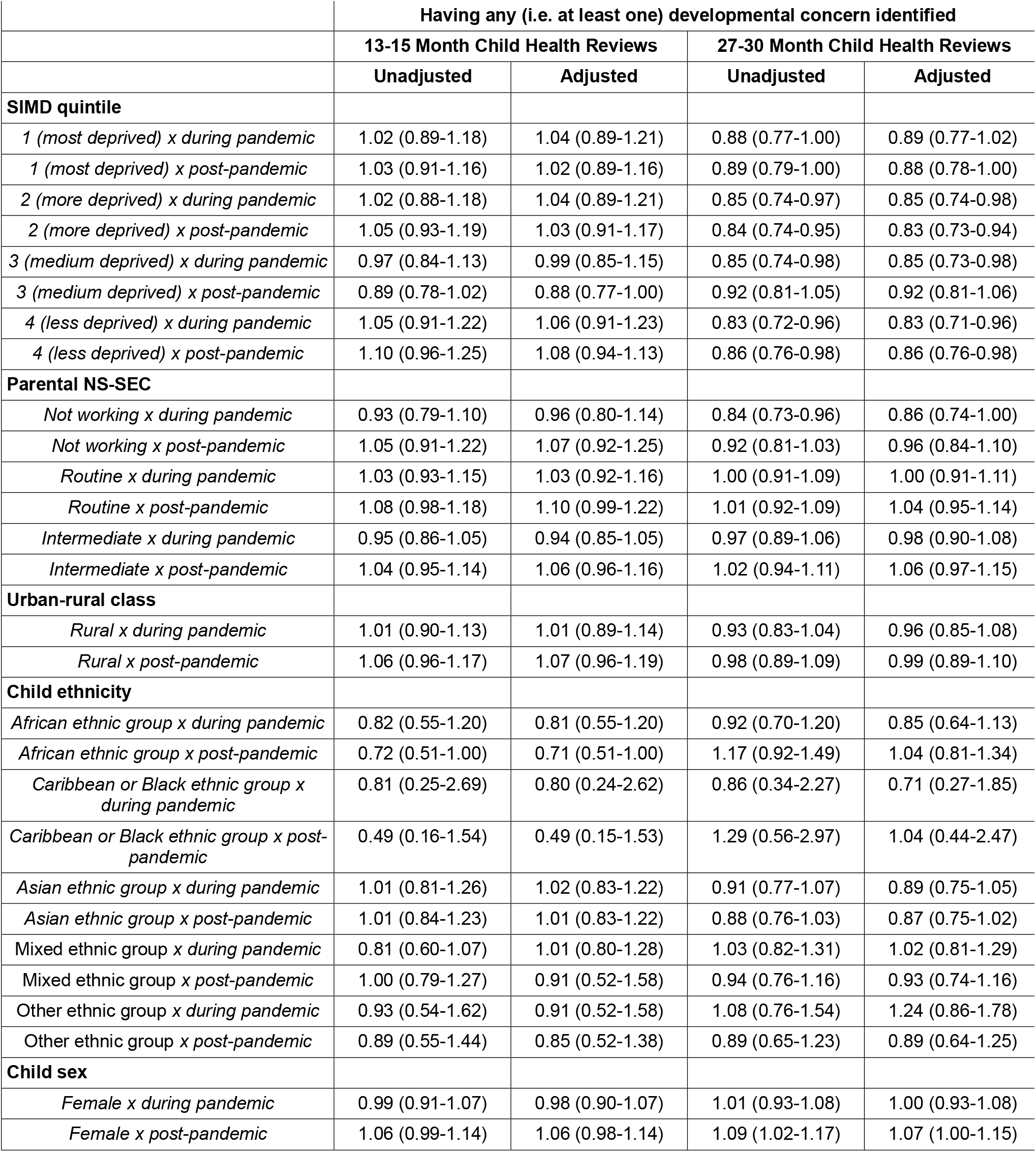
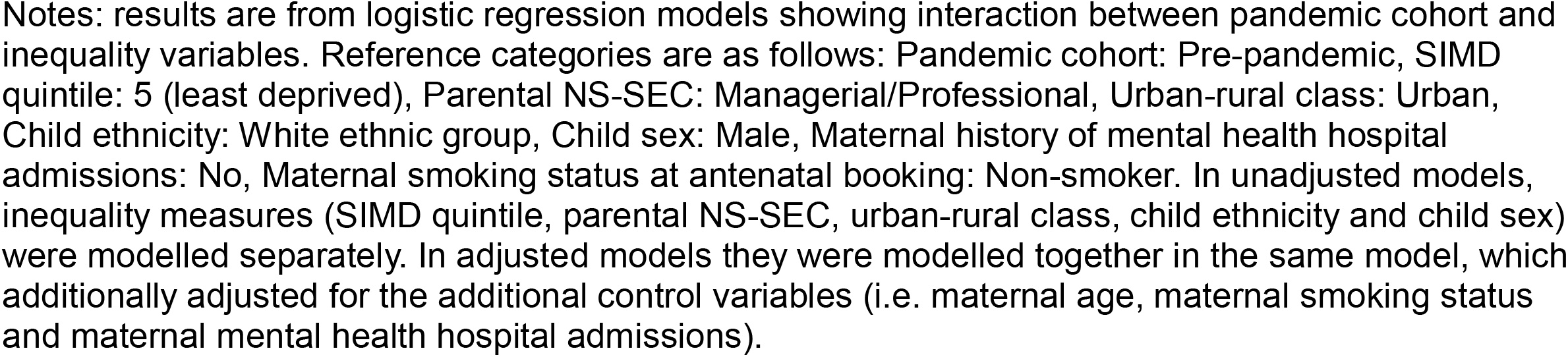
Odds ratios (95% confidence intervals) showing interaction between pandemic cohort and inequality variables.

|  | Having any (i.e. at least one) developmental concern identified |  |  |  |
| --- | --- | --- | --- | --- |
|  | 13-15 Month Child Health Reviews |  | 27-30 Month Child Health Reviews |  |
|  | Unadjusted | Adjusted | Unadjusted | Adjusted |
| **SIMD quintile** |  |  |  |  |
| *1 (most deprived) x during pandemic* | 1.02 (0.89-1.18) | 1.04 (0.89-1.21) | 0.88 (0.77-1.00) | 0.89 (0.77-1.02) |
| *1 (most deprived) x post-pandemic* | 1.03 (0.91-1.16) | 1.02 (0.89-1.16) | 0.89 (0.79-1.00) | 0.88 (0.78-1.00) |
| *2 (more deprived) x during pandemic* | 1.02 (0.88-1.18) | 1.04 (0.89-1.21) | 0.85 (0.74-0.97) | 0.85 (0.74-0.98) |
| *2 (more deprived) x post-pandemic* | 1.05 (0.93-1.19) | 1.03 (0.91-1.17) | 0.84 (0.74-0.95) | 0.83 (0.73-0.94) |
| *3 (medium deprived) x during pandemic* | 0.97 (0.84-1.13) | 0.99 (0.85-1.15) | 0.85 (0.74-0.98) | 0.85 (0.73-0.98) |
| *3 (medium deprived) x post-pandemic* | 0.89 (0.78-1.02) | 0.88 (0.77-1.00) | 0.92 (0.81-1.05) | 0.92 (0.81-1.06) |
| *4 (less deprived) x during pandemic* | 1.05 (0.91-1.22) | 1.06 (0.91-1.23) | 0.83 (0.72-0.96) | 0.83 (0.71-0.96) |
| *4 (less deprived) x post-pandemic* | 1.10 (0.96-1.25) | 1.08 (0.94-1.13) | 0.86 (0.76-0.98) | 0.86 (0.76-0.98) |
| **Parental NS-SEC** |  |  |  |  |
| *Not working x during pandemic* | 0.93 (0.79-1.10) | 0.96 (0.80-1.14) | 0.84 (0.73-0.96) | 0.86 (0.74-1.00) |
| *Not working x post-pandemic* | 1.05 (0.91-1.22) | 1.07 (0.92-1.25) | 0.92 (0.81-1.03) | 0.96 (0.84-1.10) |
| *Routine x during pandemic* | 1.03 (0.93-1.15) | 1.03 (0.92-1.16) | 1.00 (0.91-1.09) | 1.00 (0.91-1.11) |
| *Routine x post-pandemic* | 1.08 (0.98-1.18) | 1.10 (0.99-1.22) | 1.01 (0.92-1.09) | 1.04 (0.95-1.14) |
| *Intermediate x during pandemic* | 0.95 (0.86-1.05) | 0.94 (0.85-1.05) | 0.97 (0.89-1.06) | 0.98 (0.90-1.08) |
| *Intermediate x post-pandemic* | 1.04 (0.95-1.14) | 1.06 (0.96-1.16) | 1.02 (0.94-1.11) | 1.06 (0.97-1.15) |
| **Urban-rural class** |  |  |  |  |
| *Rural x during pandemic* | 1.01 (0.90-1.13) | 1.01 (0.89-1.14) | 0.93 (0.83-1.04) | 0.96 (0.85-1.08) |
| *Rural x post-pandemic* | 1.06 (0.96-1.17) | 1.07 (0.96-1.19) | 0.98 (0.89-1.09) | 0.99 (0.89-1.10) |
| **Child ethnicity** |  |  |  |  |
| *African ethnic group x during pandemic* | 0.82 (0.55-1.20) | 0.81 (0.55-1.20) | 0.92 (0.70-1.20) | 0.85 (0.64-1.13) |
| *African ethnic group x post-pandemic* | 0.72 (0.51-1.00) | 0.71 (0.51-1.00) | 1.17 (0.92-1.49) | 1.04 (0.81-1.34) |
| *Caribbean or Black ethnic group x during pandemic* | 0.81 (0.25-2.69) | 0.80 (0.24-2.62) | 0.86 (0.34-2.27) | 0.71 (0.27-1.85) |
| *Caribbean or Black ethnic group x post-pandemic* | 0.49 (0.16-1.54) | 0.49 (0.15-1.53) | 1.29 (0.56-2.97) | 1.04 (0.44-2.47) |
| *Asian ethnic group x during pandemic* | 1.01 (0.81-1.26) | 1.02 (0.83-1.22) | 0.91 (0.77-1.07) | 0.89 (0.75-1.05) |
| *Asian ethnic group x post-pandemic* | 1.01 (0.84-1.23) | 1.01 (0.83-1.22) | 0.88 (0.76-1.03) | 0.87 (0.75-1.02) |
| *Mixed ethnic group x during pandemic* | 0.81 (0.60-1.07) | 1.01 (0.80-1.28) | 1.03 (0.82-1.31) | 1.02 (0.81-1.29) |
| *Mixed ethnic group x post-pandemic* | 1.00 (0.79-1.27) | 0.91 (0.52-1.58) | 0.94 (0.76-1.16) | 0.93 (0.74-1.16) |
| *Other ethnic group x during pandemic* | 0.93 (0.54-1.62) | 0.91 (0.52-1.58) | 1.08 (0.76-1.54) | 1.24 (0.86-1.78) |
| *Other ethnic group x post-pandemic* | 0.89 (0.55-1.44) | 0.85 (0.52-1.38) | 0.89 (0.65-1.23) | 0.89 (0.64-1.25) |
| **Child sex** |  |  |  |  |
| *Female x during pandemic* | 0.99 (0.91-1.07) | 0.98 (0.90-1.07) | 1.01 (0.93-1.08) | 1.00 (0.93-1.08) |
| *Female x post-pandemic* | 1.06 (0.99-1.14) | 1.06 (0.98-1.14) | 1.09 (1.02-1.17) | 1.07 (1.00-1.15) |

Notes: results are from logistic regression models showing interaction between pandemic cohort and inequality variables. Reference categories are as follows: Pandemic cohort: Pre-pandemic, SIMD quintile: 5 (least deprived), Parental NS-SEC: Managerial/Professional, Urban-rural class: Urban, Child ethnicity: White ethnic group, Child sex: Male, Maternal history of mental health hospital admissions: No, Maternal smoking status at antenatal booking: Non-smoker. In unadjusted models, inequality measures (SIMD quintile, parental NS-SEC, urban-rural class, child ethnicity and child sex) were modelled separately. In adjusted models they were modelled together in the same model, which additionally adjusted for the additional control variables (i.e. maternal age, maternal smoking status and maternal mental health hospital admissions).

Results of the sensitivity analysis excluding children from the Greater Glasgow and Clyde Health board are provided in supplementary appendix Tables A3 and A4. Results of the sensitivity analysis additionally adjusting for child gestational age at birth are provided in supplementary appendix Tables A5 and A6. Findings from both sensitivity analyses did not significantly differ from the results of the primary analysis. This suggests that the results of our primary analysis were not driven by issues relating to data issues in Greater Glasgow and Clyde, including later review implementation and incomplete data on assessment of problem-solving developmental concerns in early 2019, and held with and without accounting for the potentially mediating effect of gestational age.

## Discussion

Our population-based retrospective cohort study, conducted as part of the wider CHILDS project, used linked administrative health data to investigate developmental inequalities in Scotland before, during and after the COVID-19 pandemic. Our findings suggest that substantial developmental inequalities persist, with children who are male, from deprived or urban areas, with parents in lower socioeconomic occupations and/or from African or Asian ethnic groups being more likely to have developmental concerns. However, these developmental inequalities did not widen during the pandemic. Across almost all measures, interaction analyses showed no significant change in inequalities during or after the pandemic compared to the pre-pandemic period, and some SIMD-based inequalities showed modest narrowing. This aligns with finding from studies of older children from the UK and Germany [25, 26], but differs from the findings from the majority of quantitative studies from other countries, which reported widening disparities [17-24].

Consistent with previous research [13], we observed an overall increase child developmental concerns during the pandemic. There were concerns that this rise would disproportionately impact those from low-income households, urban areas or minority ethnic groups. However, reassuringly, this does not appear to have been the case as, although the proportion of children with developmental concerns increased overall, pre-existing inequalities were not exacerbated. This suggests that universal systems in Scotland such as the National Health Service may have buffered some of the PHSM impacts on developmental inequalities seen in other countries that do not have universal provisions to the same extent. Specific policies introduced during the pandemic, such as job retention schemes set up to protect families against income losses, and childcare hubs set up to early learning and childcare to the children of key workers and those deemed vulnerable, may also have buffered against the widening of inequalities.

Nevertheless, although inequalities did not widen, overall levels of developmental concerns did increase during the pandemic, and the scale of persistent developmental inequalities remains striking. At age 27-30 months, children from the most deprived areas had approximately twice the odds of developmental concerns compared with those from the least deprived areas, even after adjustment for socioeconomic factors and maternal covariates. Similarly, children whose parents were not working had more than twice the odds of concerns compared with those whose parents were in managerial or professional occupations.

These findings are highly relevant to the Scottish Government’s target of reducing the proportion of children with 27-30 month developmental concerns by 25% by 2030. Achieving this will require addressing underlying socioeconomic inequalities. Recent policies, including the Scottish Child Payment (a weekly payment supporting families that is received for every child under 16), Scotland’s expanded entitlement to funded early learning and childcare (from 600 to 1,140 per year for all three and four year olds and eligible two year olds), and the UK-wide removal of the Universal Credit 2-child limit (which restricted UK welfare support to two children per household only), may aid this. However, further interventions aimed at reducing child poverty, redistributing resources and promoting child health are likely to be required to further reduce socioeconomic developmental inequalities. This may include creating a more supportive environment within the UK welfare system (e.g. by replacing ‘work first’ and benefit sanctions with a more ‘rights-based’ approach), expanding benefit maximisation schemes and Financial Inclusion Support Officers (FISOs), trialling cash transfer schemes (similar to the ‘Baby’s First Years Project’ in the US), new Sure Start policies, and health interventions from pre-conception, pregnancy and infancy to reduce premature birth (e.g. smoking cessation support) and increase breastfeeding. Addressing socioeconomic inequalities at their roots in the early years has potential to provide lasting benefits throughout children’s lifetimes.

In addition to socioeconomic inequalities, our findings also highlight the significantly higher rates of developmental concerns in boys compared to girls in Scotland. At 27-30 months, girls had approximately 60% lower odds of concerns than boys. This pattern is also observed internationally, and is particularly marked in language and socio-emotional domains [33]. The mechanisms underlying this disparity remain unclear, and may reflect biological factors, differences in developmental trajectories, or potential measurement biases in existing assessment tools.

The key strength of our analysis was its large, population-based sample, enabling robust examination of developmental inequalities across three distinct pandemic periods. However, there are some limitations to note. First, although our study’s coverage was high (we estimate that over 80% of all children in Scotland aged 13-15 months or 27-30 months during our analysis period were included), those who did not receive reviews or were incompletely assessed were excluded. Second, parental NS-SEC was derived from child birth records and therefore does not capture changes in parental occupation post-birth. Third, during the early periods of COVID-19 PHSM, some health reviews were conducted remotely which may have influenced detection of developmental concerns. Fourth, health-board level variation in review implementation and data completeness, particularly in Greater Glasgow and Clyde in early 2019, may have affected early-period data; however, sensitivity analyses excluding this region suggests this did not influence our results. Finally, our analysis did not include data on language spoken in the home. Some inequality variables may be proxies of not having English as a first language.

Despite these limitations, our study provides important new data on developmental inequalities in Scotland before, during and after the COVID-19 pandemic. Although the pandemic was associated with an overall rise in developmental concerns, it did not lead to systematic widening of pre-existing inequalities. While this is reassuring, the persistence and magnitude of pre-existing socioeconomic disparities underscore the need for interventions to reduce inequalities and meet national child development targets.

## Data Availability

The administrative health datasets used for this study cannot be publicly shared by the authors. They can however be accessed via successfully applying to the NHS Scotland Public Benefit and Privacy Panel for Health and Social Care (HSC-PBPP).

## Contributors

IH: conceptualisation, data curation, project administration, formal analysis, methodology, writing - original draft. LM: conceptualisation, methodology, funding acquisition, writing - review and editing. AM: conceptualisation, methodology, funding acquisition, writing - review and editing. JK: conceptualisation, methodology, writing - review and editing. KO: conceptualisation, data curation - review and verification, formal analysis - review and verification, writing - review and editing. JPB: conceptualisation, methodology, writing – review and editing. MVL: conceptualisation, writing - review and editing. PW: conceptualisation, writing - review and editing. RW: conceptualisation, methodology, funding acquisition, writing - review and editing. BA: conceptualisation, methodology, supervision, project administration, funding acquisition, methodology, writing - review and editing. IH and KO accessed and verified the raw data.

## Acknowledgements

This study was funded by the Economic and Social Research Council (ES/W001519/1). BA was also supported by the EU Horizon 2020 research and innovation programme under the Marie Skłodowska-Curie grant agreement number 813546, and Baily Thomas Charitable Fund (TRUST/VC/AC/SG/469207686). We would like to thank and acknowledge the eDRIS team at Public Health Scotland for their support in obtaining approvals, the provisioning and linking of data, and facilitating access to the data in Scotland’s National Safe Haven. All other authors could access the raw data if they wished, provided that they completed information governance training in order to be granted access to the data in Scotland’s National Safe Haven by the Electronic Data Research and Innovation Service (eDRIS) team at public health Scotland. All authors had final responsibility for the decision to submit for publication.

## Data sharing statement

The administrative health datasets used for this study cannot be publicly shared by the authors, but can be accessed via successfully applying to the NHS Scotland Public Benefit and Privacy Panel for Health and Social Care (HSC-PBPP). The authors of the present study were supported in applying for approval from HSC-PBPP by the eDRIS team at Public Health Scotland. eDRIS also facilitated access to the data via Scotland’s National Safe Haven.

## Patient and public involvement

When applying for funding for the wider CHILDS study, mothers with lived experience of giving birth during the pandemic fed into our research aims and shared their experience of child health reviews and which outcomes were most relevant to their families.

## References

1. World Health Organization, United Nations Children’s Fund, and W.B. Group, Nurturing care for early childhood development: a framework for helping children survive and thrive to transform health and human potential. 2018.

2. Pearce, A., R. Dundas, M. Whitehead, and D. Taylor-Robinson, Pathways to inequalities in child health. Arch Dis Child, 2019. 104(10): p. 998–1003.

3. Webster, E.M., The Impact of Adverse Childhood Experiences on Health and Development in Young Children. Glob Pediatr Health, 2022. 9: p. 2333794x221078708.

4. Wallerich, L., A. Fillol, A. Rivadeneyra, S. Vandentorren, J. Wittwer, and L. Cambon, Environment and child well-being: A scoping review of reviews to guide policies. Health Promot Perspect, 2023. 13(3): p. 168–182.

5. Cattan, S., E. Fitzsimons, A. Goodman, A. Phimister, G.B. Ploubidis, and J. Wertz, Early childhood inequalities. Oxford Open Economics, 2024. 3(Supplement_1): p. i711–i740.

6. Midouhas, E. and L. Platt, Rural–urban area of residence and trajectories of childrenfZs behavior in England. Health & Place, 2014. 30: p. 226–233.

7. Jamieson, L., P. Bradshaw, and R. Ormiston, Growing Up In Scotland Study: Growing Up In Rural Scotland. 2008 Education Analytical Services, Scottish Government: Edinburgh.

8. Scottish Parliament Information Centre. Timeline of Coronavirus (COVID-19) in Scotland. 2023; Available from: https://spice-spotlight.scot/2023/05/10/timeline-of-coronavirus-covid-19-in-scotland/.

9. Mahase, E., Covid-19: Lockdowns and masks helped reduce transmission, expert group finds. BMJ, 2023. 382: p. 1959.

10. Fox, L., C. Bowyer-Crane, A.A. Lambrechts, C. Manzoni, D. Nielsen, and L. Tracey. Mitigating Impacts of COVID-19 in the Early Years - Rapid Evidence Review. 2021; Available from: https://www.york.ac.uk/education/news-events/news/2021/early-years-covid19-report-released/.

11. Green, M.J., A. Pearce, A. Parkes, E. Robertson, and S.V. Katikireddi, Pre-school childcare and inequalities in child development. SSM Popul Health, 2021. 14: p. 100776.

12. Hardie, I., L. Marryat, A. Murray, et al., Early childhood developmental concerns following SARS-CoV-2 infection and COVID-19 vaccination during pregnancy: a Scottish population-level retrospective cohort study. The Lancet Child & Adolescent Health, 2025. 9(3): p. 162–171.

13. Hardie, I., L. Marryat, A. Murray, et al., COVID-19 public health and social measures (PHSM) and early childhood developmental concerns in Scotland: an interrupted time series analysis. Lancet Reg Health Eur, 2026. 60: p. 101525.

14. Alcon, S., S. Shen, H.N. Wong, et al., Effects of the COVID-19 Pandemic on Early Childhood Development and Mental Health: A Systematic Review and Meta-Analysis of Comparative Studies. Psychol Int, 2024. 6(4): p. 986–1012.

15. Public Health Scotland, The impact of COVID-19 on children and young people in Scotland: 2 to 4 year olds. 2020.

16. Pajek, J., K. Mancini, and M. Murray, COVID-19 and children’s behavioral health: An overview. Current Problems in Pediatric and Adolescent Health Care, 2023. 53(10): p. 101491.

17. Fung, P., T. St. Pierre, M. Raja, and E.K. Johnson, Infants’ and toddlers’ language development during the pandemic: Socioeconomic status mattered. Journal of Experimental Child Psychology, 2023. 236: p. 105744.

18. Scott, R.M., G. Nguyentran, and J.Z. Sullivan, The COVID-19 pandemic and social cognitive outcomes in early childhood. Scientific Reports, 2024. 14(1): p. 28939.

19. Sharp, H., N. Wright, L. Bozicevic, et al., Inequalities in COVID-19 impact on preschool mental health in India: key moderators of adverse outcome. BMJ Public Health, 2024. 2(2): p. e001209.

20. González, M., T. Loose, M. Liz, et al., School readiness losses during the COVID-19 outbreak. A comparison of two cohorts of young children. Child Development, 2022. 93(4): p. 910–924.

21. Romem, S., M. Katusic, C.-I. Wi, R. Hentz, and B.A. Lynch, A retrospective cohort study analyzing the changes in early childhood development during the COVID-19 pandemic. Early Human Development, 2024. 192: p. 105991.

22. Hoffmann, S., M. Tschorn, and J. Spallek, Social inequalities in early childhood language development during the COVID-19 pandemic: a descriptive study with data from three consecutive school entry surveys in Germany. International Journal for Equity in Health, 2024. 23(1): p. 2.

23. Lee, K.-S., Y.Y. Choi, Y.S. Kim, Y. Kim, M.-H. Kim, and N. Lee, Association between the COVID-19 pandemic and childhood development aged 30 to 36 months in South Korea, based on the National health screening program for infants and children database. BMC Public Health, 2024. 24(1): p. 989.

24. Hamadani, J.D., S.A. Bhuiyan, M.I. Hasan, et al., The effect of exposure to the COVID-19 pandemic on nutritional status and cognitive, motor, and behavioural development among children aged 20 months in rural Bangladesh: A repeated cross-section study between 2020 and 2022. PLoS One, 2025. 20(3): p. e0309836.

25. Rigó, M. and S. Weyers, Child Motor Development before and after the COVID-19 Pandemic: Are There Social Inequalities? Children, 2024. 11(8): p. 936.

26. Miall, N., A. Pearce, J.C. Moore, M. Benzeval, and M.J. Green, Inequalities in children’s mental health before and during the COVID-19 pandemic: findings from the UK Household Longitudinal Study. Journal of Epidemiology and Community Health, 2023: p. jech-2022-220188.

27. Information Services Division Scotland. SMR04 - Mental Health Inpatient and Day Case. SMR Datasets 2022; Available from: https://www.ndc.scot.nhs.uk/Data-Dictionary/SMR-Datasets/SMR04-Mental-Health-Inpatient-and-Day-Case/.

28. Stock, S.J., J. Carruthers, C. Denny, et al., Cohort Profile: The COVID-19 in Pregnancy in Scotland (COPS) dynamic cohort of pregnant women to assess effects of viral and vaccine exposures on pregnancy. International Journal of Epidemiology, 2022. 51(5): p. e245–e255.

29. Scottish Government. Universal Health Visiting Pathway in Scotland: Pre-Birth to Pre-School. 2015 01/02/2023]; Available from: https://www.gov.scot/binaries/content/documents/govscot/publications/advice-and-guidance/2015/10/universal-health-visiting-pathway-scotland-pre-birth-pre-school/documents/00487884-pdf/00487884-pdf/govscot%3Adocument/00487884.pdf.

30. Public Health Scotland. National Datasets: National Records of Scotland (NRS) Births, Stillbirths and Infant Deaths. 2025; Available from: https://publichealthscotland.scot/resources-and-tools/health-intelligence-and-data-management/national-data-catalogue/national-datasets/search-the-datasets/national-records-of-scotland-nrs-births-stillbirths-and-infant-deaths/.

31. Public Health Scotland. Child health pre-school review coverage: 2023 to 2024. 2025; Available from: https://publichealthscotland.scot/publications/child-health-pre-school-review-coverage/child-health-pre-school-review-coverage-2023-to-2024/.

32. Scottish Government. Scottish Government Urban Rural Classification 2020. 2022; Available from: https://www.gov.scot/publications/scottish-government-urban-rural-classification-2020/documents/.

33. Bando, R., F. Lopez-Boo, L. Fernald, P. Gertler, and S. Reynolds, Gender Differences in Early Child Development: Evidence from Large-Scale Studies of Very Young Children in Nine Countries. Journal of Economics, Race, and Policy, 2024. 7(2): p. 82–92.

