## Supplementary Appendix for "Inequalities in early childhood developmental concerns before, during and after the COVID-19 pandemic in Scotland: a retrospective cohort study"

### **Table A1. The RECORD statement – checklist of items, extended from the STROBE statement, that should be reported in observational studies using routinely collected health data.**

|  | **Item No.** | **STROBE items** | **Location in manuscript where items are reported** | **RECORD items** | **Location in manuscript where items are reported** |
| --- | --- | --- | --- | --- | --- |
| **Title and abstract** | | | | | |
|  | 1 | (a) Indicate the study’s design with a commonly used term in the title or the abstract (b) Provide in the abstract an informative and balanced summary of what was done and what was found | Title and abstract | RECORD 1.1: The type of data used should be specified in the title or abstract. When possible, the name of the databases used should be included.  RECORD 1.2: If applicable, the geographic region and timeframe within which the study took place should be reported in the title or abstract.  RECORD 1.3: If linkage between databases was conducted for the study, this should be clearly stated in the title or abstract. | Title  Title (p1) and abstract  Abstract |
| Background rationale | 2 | Explain the scientific background and rationale for the investigation being reported | Abstract |  |  |
| Objectives | 3 | State specific objectives, including any prespecified hypotheses | Abstract |  |  |
| Study Design | 4 | Present key elements of study design early in the paper | Study design and participants section |  |  |
| Setting | 5 | Describe the setting, locations, and relevant dates, including periods of recruitment, exposure, follow-up, and data collection | Study design and participants section |  |  |
| Participants | 6 | *(a) Cohort study* - Give the eligibility criteria, and the sources and methods of selection of participants. Describe methods of follow-up  *Case-control study* - Give the eligibility criteria, and the sources and methods of case ascertainment and control selection. Give the rationale for the choice of cases and controls  *Cross-sectional study* - Give the eligibility criteria, and the sources and methods of selection of participants  *(b) Cohort study* - For matched studies, give matching criteria and number of exposed and unexposed  *Case-control study* - For matched studies, give matching criteria and the number of controls per case | Study design and participants section | RECORD 6.1: The methods of study population selection (such as codes or algorithms used to identify subjects) should be listed in detail. If this is not possible, an explanation should be provided.  RECORD 6.2: Any validation studies of the codes or algorithms used to select the population should be referenced. If validation was conducted for this study and not published elsewhere, detailed methods and results should be provided.  RECORD 6.3: If the study involved linkage of databases, consider use of a flow diagram or other graphical display to demonstrate the data linkage process, including the number of individuals with linked data at each stage. | Study design and participants section  In text in study design and participants section |
| Variables | 7 | Clearly define all outcomes, exposures, predictors, potential confounders, and effect modifiers. Give diagnostic criteria, if applicable. | Procedures section | RECORD 7.1: A complete list of codes and algorithms used to classify exposures, outcomes, confounders, and effect modifiers should be provided. If these cannot be reported, an explanation should be provided. | Procedures section |
| Data sources/ measurement | 8 | For each variable of interest, give sources of data and details of methods of assessment (measurement).  Describe comparability of assessment methods if there is more than one group | Procedures section |  |  |
| Bias | 9 | Describe any efforts to address potential sources of bias | Statistical analysis section (p8-9) |  |  |
| Study size | 10 | Explain how the study size was arrived at | Study design and participants section |  |  |
| Quantitative variables | 11 | Explain how quantitative variables were handled in the analyses. If applicable, describe which groupings were chosen, and why | Procedures section |  |  |
| Statistical methods | 12 | (a) Describe all statistical methods, including those used to control for confounding  (b) Describe any methods used to examine subgroups and interactions  (c) Explain how missing data were addressed  (d) *Cohort study* - If applicable, explain how loss to follow-up was addressed  *Case-control study* - If applicable, explain how matching of cases and controls was addressed  *Cross-sectional study* - If applicable, describe analytical methods taking account of sampling strategy  (e) Describe any sensitivity analyses | Statistical analysis section |  |  |
| Data access and cleaning methods |  | .. |  | RECORD 12.1: Authors should describe the extent to which the investigators had access to the database population used to create the study population.  RECORD 12.2: Authors should provide information on the data cleaning methods used in the study. | Study design and participants section, Procedures section and Statistical analysis section |
| Linkage |  | .. |  | RECORD 12.3: State whether the study included person-level, institutional-level, or other data linkage across two or more databases. The methods of linkage and methods of linkage quality evaluation should be provided. | Study design and participants section |
| Participants | 13 | (a) Report the numbers of individuals at each stage of the study (*e.g.*, numbers potentially eligible, examined for eligibility, confirmed eligible, included in the study, completing follow-up, and analysed)  (b) Give reasons for non-participation at each stage.  (c) Consider use of a flow diagram | Study design and participants section | RECORD 13.1: Describe in detail the selection of the persons included in the study (*i.e.,* study population selection) including filtering based on data quality, data availability and linkage. The selection of included persons can be described in the text and/or by means of the study flow diagram. | Study design and participants section |
| Descriptive data | 14 | (a) Give characteristics of study participants (*e.g.*, demographic, clinical, social) and information on exposures and potential confounders  (b) Indicate the number of participants with missing data for each variable of interest  (c) *Cohort study* - summarise follow-up time (*e.g.*, average and total amount) | Results section |  |  |
| Outcome data | 15 | *Cohort study* - Report numbers of outcome events or summary measures over time  *Case-control study* - Report numbers in each exposure category, or summary measures of exposure  *Cross-sectional study* - Report numbers of outcome events or summary measures |  |  |  |
| Main results | 16 | (a) Give unadjusted estimates and, if applicable, confounder-adjusted estimates and their precision (e.g., 95% confidence interval). Make clear which confounders were adjusted for and why they were included  (b) Report category boundaries when continuous variables were categorized  (c) If relevant, consider translating estimates of relative risk into absolute risk for a meaningful time period | Results section |  |  |
| Other analyses | 17 | Report other analyses done—e.g., analyses of subgroups and interactions, and sensitivity analyses | Results section |  |  |
| Key results | 18 | Summarise key results with reference to study objectives | Results section and discussion section |  |  |
| Limitations | 19 | Discuss limitations of the study, taking into account sources of potential bias or imprecision. Discuss both direction and magnitude of any potential bias | Discussion section | RECORD 19.1: Discuss the implications of using data that were not created or collected to answer the specific research question(s). Include discussion of misclassification bias, unmeasured confounding, missing data, and changing eligibility over time, as they pertain to the study being reported. | Discussion section |
| Interpretation | 20 | Give a cautious overall interpretation of results considering objectives, limitations, multiplicity of analyses, results from similar studies, and other relevant evidence | Discussion section |  |  |
| Generalisability | 21 | Discuss the generalisability (external validity) of the study results | Discussion section |  |  |
| Funding | 22 | Give the source of funding and the role of the funders for the present study and, if applicable, for the original study on which the present article is based | Acknowledgements section |  |  |
| Accessibility of protocol, raw data, and programming code |  | .. |  | RECORD 22.1: Authors should provide information on how to access any supplemental information such as the study protocol, raw data, or programming code. | Data sharing statement |

*Reference: Benchimol EI, Smeeth L, Guttmann A, Harron K, Moher D, Petersen I, Sørensen HT, von Elm E, Langan SM, the RECORD Working Committee. The REporting of studies Conducted using Observational Routinely-collected health Data (RECORD) Statement. *PLoS Medicine* 2015; in press. *Checklist is protected under Creative Commons Attribution ([CC BY](http://creativecommons.org/licenses/by/4.0/)) license.

**Figure A1. Unadjusted odds ratios and 95% confidence intervals showing inequalities in odds of developmental concerns at age 13-15 months before, during and after the COVID-19 pandemic**

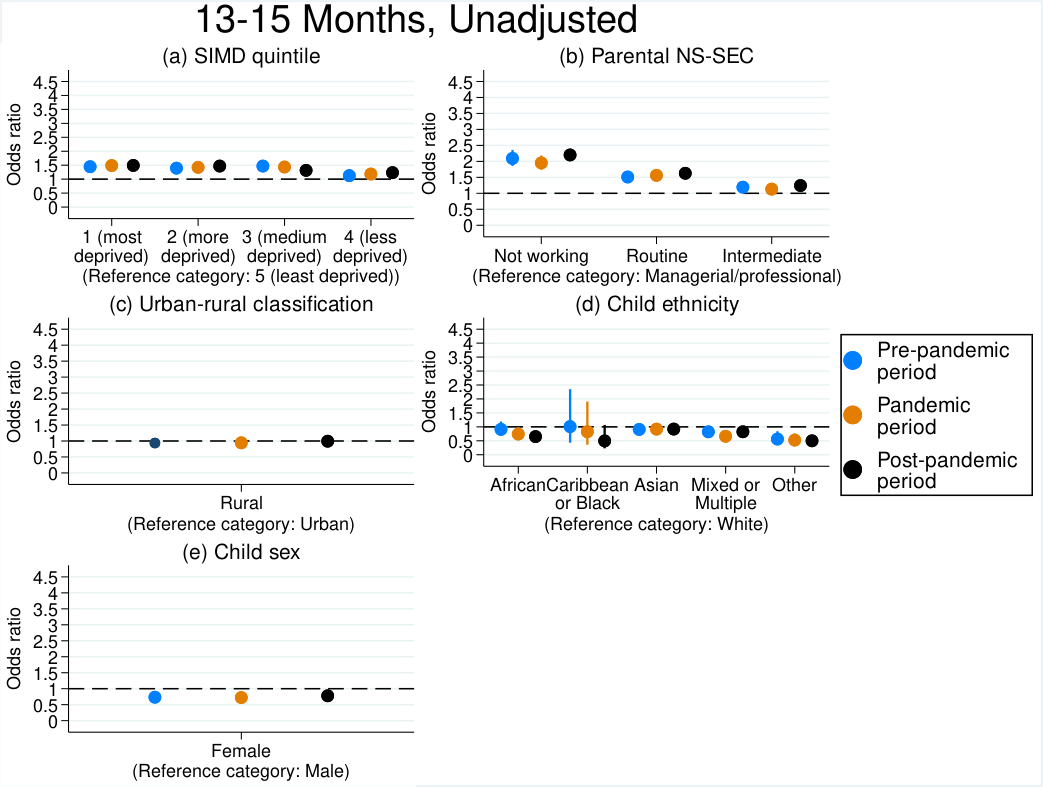

Notes: odds ratios are shown by point estimates and 95% confidence intervals are shown by vertical bars. The dotted line indicates no difference in odds ratios compared to reference groups, whilst higher odds ratios above this indicate increased odds of developmental concerns compared to reference groups. Models were conducted separately for the pre-pandemic cohort, pandemic cohort and post-pandemic cohort, and odds ratios can be compared to see how inequalities differed over time between cohorts.

**Figure A2. Unadjusted odds ratios and 95% confidence intervals showing inequalities in odds of developmental concerns at age 27-30 months before, during and after the COVID-19 pandemic**

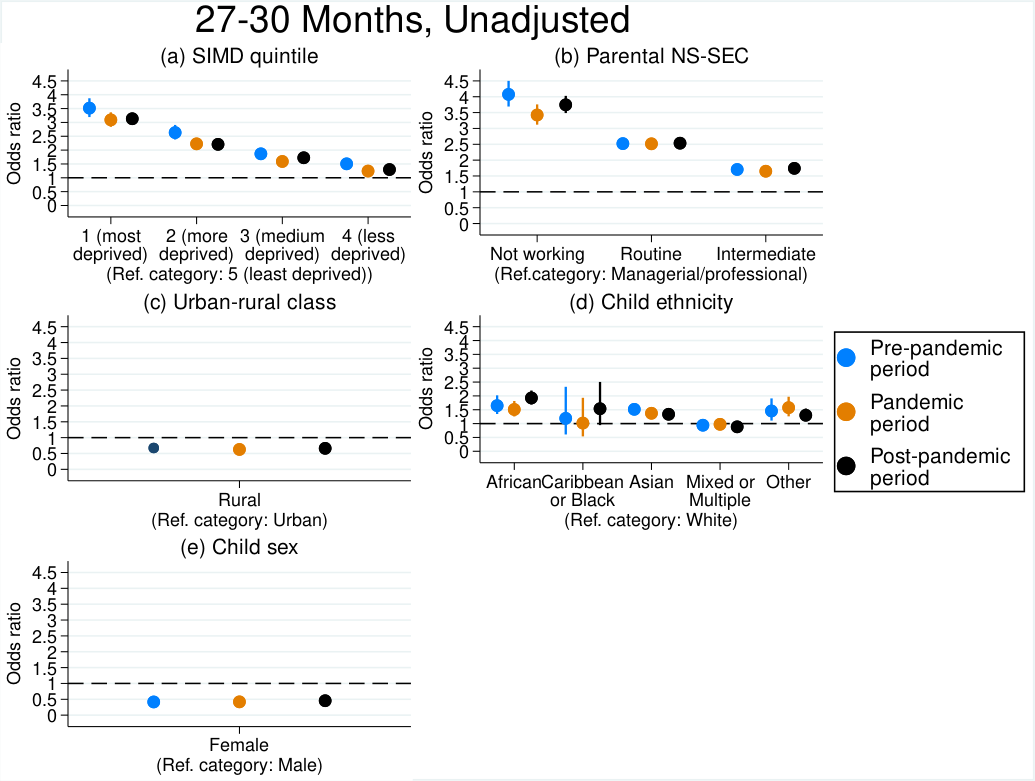

Notes: odds ratios are shown by point estimates and 95% confidence intervals are shown by vertical bars. The dotted line indicates no difference in odds ratios compared to reference groups, whilst higher odds ratios above this indicate increased odds of developmental concerns compared to reference groups. Models were conducted separately for the pre-pandemic cohort, pandemic cohort and post-pandemic cohort, and odds ratios can be compared to see how inequalities differed over time between cohorts.

**Table A2. Odds ratios (95% confidence intervals) showing inequalities in odds of developmental concerns before, during and after the COVID-19 pandemic**

|  | **Having any (i.e. at least one) developmental concern identified** | | | | | | | | | | | |
| --- | --- | --- | --- | --- | --- | --- | --- | --- | --- | --- | --- | --- |
|  | **13-15 Month Child Health Reviews** | | | | | | **27-30 Month Child Health Reviews** | | | | | |
|  | **Pre-pandemic**  **period** | | **Pandemic**  **period** | | **Post-pandemic period** | | **Pre-pandemic**  **period** | | **Pandemic**  **period** | | **Post-pandemic period** | |
|  | **Un-adjusted** | **Adjusted** | **Un-adjusted** | **Adjusted** | **Un-adjusted** | **Adjusted** | **Un-adjusted** | **Adjusted** | **Un-adjusted** | **Adjusted** | **Un-adjusted** | **Adjusted** |
| **SIMD quintile** |  |  |  |  |  |  |  |  |  |  |  |  |
| *1 (most deprived)* | 1.45  (1.31-1.60) | 1.16  (1.04-1.29) | 1.49  (1.35-1.64) | 1.17  (1.05-1.30) | 1.49  (1.39-1.60) | 1.13  (1.05-1.22) | 3.52  (3.19-3.87) | 2.27  (2.04-2.52) | 3.09  (2.84-3.36) | 2.01  (1.84-2.21) | 3.13  (2.93-3.35) | 1.97  (1.83-2.12) |
| *2 (more deprived)* | 1.39  (1.26-1.54) | 1.20  (1.08-1.33) | 1.42  (1.29-1.57) | 1.22  (1.10-1.36) | 1.47  (1.37-1.58) | 1.20  (1.11-1.29) | 2.63  (2.38-2.91) | 1.95  (1.76-2.17) | 2.23  (2.04-2.43) | 1.65  (1.51-1.82) | 2.21  (2.06-2.37) | 1.61  (1.49-1.73) |
| *3 (medium deprived)* | 1.47  (1.33-1.63) | 1.35  (1.21-1.50) | 1.44  (1.30-1.59) | 1.32  (1.19-1.47) | 1.32  (1.22-1.42) | 1.16  (1.08-1.26) | 1.87  (1.68-2.08) | 1.60  (1.44-1.79) | 1.59  (1.45-1.74) | 1.36  (1.24-1.51) | 1.72  (1.60-1.86) | 1.47  (1.36-1.59) |
| *4 (less deprived)* | 1.13  (1.01-1.25) | 1.09  (0.98-1.22) | 1.18  (1.07-1.31) | 1.15  (1.04-1.28) | 1.23  (1.15-1.32) | 1.16  (1.08-1.26) | 1.50  (1.35-1.68) | 1.44  (1.29-1.61) | 1.24  (1.13-1.37) | 1.19  (1.08-1.32) | 1.30  (1.20-1.40) | 1.24  (1.14-1.33) |
| **Parental NSSEC** |  |  |  |  |  |  |  |  |  |  |  |  |
| *Not working* | 2.09  (1.86-2.36) | 1.96  (1.72-2.25) | 1.94  (1.74-2.18) | 1.75  (1.54-1.99) | 2.20  (2.02-2.39) | 1.91  (1.74-2.10) | 4.07  (3.69-4.50) | 2.38  (2.12-2.66) | 3.42  (3.12-3.76) | 2.04  (1.84-2.27) | 3.75  (3.48-4.03) | 2.22  (2.04-2.41) |
| *Routine* | 1.51  (1.40-1.63) | 1.40  (1.29-1.53) | 1.56  (1.45-1.68) | 1.38  (1.27-1.51) | 1.63  (1.54-1.72) | 1.44  (1.35-1.54) | 2.52  (2.36-2.70) | 1.71  (1.58-1.84) | 2.52  (2.37-2.68) | 1.71  (1.59-1.83) | 2.53  (2.41-2.66) | 1.74  (1.64-1.83) |
| *Intermediate* | 1.19  (1.11-1.28) | 1.15  (1.06-1.24) | 1.13  (1.05-1.21) | 1.06  (0.98-1.14) | 1.24  (1.18-1.31) | 1.17  (1.10-1.23) | 1.71  (1.60-1.82) | 1.35  (1.26-1.45) | 1.65  (1.55-1.75) | 1.33  (1.25-1.42) | 1.74  (1.66-1.82) | 1.41  (1.35-1.49) |
| **Urban-rural class** |  |  |  |  |  |  |  |  |  |  |  |  |
| *Rural* | 0.94  (0.86-1.02) | 0.95  (0.87-1.04) | 0.94  (0.87-1.02) | 0.96  (0.89-1.05) | 0.99  (0.94-1.05) | 1.03  (0.97-1.09) | 0.67  (0.62-0.73) | 0.81  (0.75-0.89) | 0.63  (0.58-0.68) | 0.78  (0.72-0.85) | 0.66  (0.63-0.70) | 0.81  (0.76-0.86) |
| **Child ethnicity** |  |  |  |  |  |  |  |  |  |  |  |  |
| *African ethnic group* | 0.91  (0.70-1.19) | 0.83  (0.63-1.09) | 0.74  (0.56-0.98) | 0.71  (0.54-0.94) | 0.65  (0.54-0.79) | 0.64  (0.52-0.77) | 1.65  (1.35-2.02) | 1.55  (1.25-1.91) | 1.51  (1.26-1.81) | 1.31  (1.08-1.59) | 1.93  (1.69-2.19) | 1.67  (1.45-1.91) |
| *Caribbean or Black ethnic group* | 1.00  (0.43-2.35) | 0.96  (0.41-2.24) | 0.83  (0.36-1.91) | 0.80  (0.34-1.85) | 0.50  (0.23-1.07) | 0.50  (0.23-1.07) | 1.19  (0.60-2.33) | 1.39  (0.69-2.79) | 1.02  (0.54-1.93) | 1.00  (0.52-1.92) | 1.54  (0.94-2.51) | 1.49  (0.90-2.48) |
| *Asian ethnic group* | 0.91  (0.77-1.07) | 0.95  (0.81-1.12) | 0.92  (0.79-1.06) | 1.01  (0.87-1.17) | 0.92  (0.83-1.02) | 1.00  (0.90-1.11) | 1.52  (1.34-1.72) | 1.71  (1.50-1.95) | 1.37  (1.23-1.53) | 1.52  (1.35-1.71) | 1.34  (1.23-1.46) | 1.53  (1.40-1.67) |
| Mixed ethnic group | 0.82  (0.68-1.00) | 0.83  (0.68-1.01) | 0.66  (0.54-0.81) | 0.68  (0.55-0.83) | 0.82  (0.72-0.94) | 0.86  (0.75-0.98) | 0.94  (0.79-1.13) | 1.02  (0.85-1.22) | 0.97  (0.84-1.13) | 1.03  (0.89-1.21) | 0.88  (0.78-1.00) | 0.95  (0.84-1.08) |
| Other ethnic group | 0.56  (0.38-0.84) | 0.49  (0.33-0.74) | 0.52  (0.36-0.77) | 0.47  (0.32-0.69) | 0.50  (0.39-0.65) | 0.45  (0.34-0.58) | 1.46  (1.11-1.91) | 1.05  (0.79-1.39) | 1.58  (1.26-1.96) | 1.31  (1.04-1.66) | 1.30  (1.09-1.55) | 0.98  (0.82-1.17) |
| **Child sex** |  |  |  |  |  |  |  |  |  |  |  |  |
| *Female* | 0.73  (0.69-0.78) | 0.73  (0.69-0.78) | 0.72  (0.68-0.77) | 0.72  (0.68-0.76) | 0.78  (0.74-0.81) | 0.77  (0.74-0.81) | 0.42  (0.39-0.44) | 0.41  (0.38-0.43) | 0.42  (0.40-0.44) | 0.41  (0.39-0.43) | 0.46  (0.44-0.47) | 0.44  (0.42-0.45) |
| ***Maternal age, years*** |  | 1.01  (1.01-1.02) |  | 1.01  (1.00-1.02) |  | 1.00  (1.00-1.01) |  | 0.99  (0.98-0.99) |  | 0.99  (0.98-1.00) |  | 0.99  (0.98-0.99) |
| **Maternal history of mental health hospital admissions** |  |  |  |  |  |  |  |  |  |  |  |  |
| *Yes* |  | 1.62  (1.37-1.92) |  | 1.61  (1.35-1.92) |  | 1.57  (1.36-1.80) |  | 1.50  (1.29-1.75) |  | 1.51  (1.30-1.74) |  | 1.35  (1.19-1.53) |
| **Maternal smoking status at antenatal booking** |  |  |  |  |  |  |  |  |  |  |  |  |
| *Ex-smoker* |  | 1.06  (0.98-1.15) |  | 1.10  (1.02-1.19) |  | 1.12  (1.06-1.18) |  | 1.15  (1.07-1.24) |  | 1.09  (1.02-1.17) |  | 1.19  (1.13-1.25) |
| *Current smoker* |  | 1.32  (1.21-1.44) |  | 1.52  (1.40-1.66) |  | 1.53  (1.43-1.63) |  | 1.44  (1.34-1.55) |  | 1.56  (1.46-1.67) |  | 1.62  (1.53-1.71) |
| *Unknown* |  | 1.46  (1.23-1.74) |  | 1.35  (1.07-1.71) |  | 0.83  (0.56-1.21) |  | 1.47  (1.20-1.79) |  | 1.27  (1.09-1.48) |  | 1.13  (0.93-1.36) |

Notes: results are from logistic regression models showing associations between each inequality measure and odds of children having any (i.e. at least one) developmental concerns. Reference categories are as follows: SIMD quintile: 5 (least deprived), Parental NSSEC: Managerial/Professional, Urban-rural class: Urban, Child ethnicity: White ethnic group, Child sex: Male, Maternal history of mental health hospital admissions: No, Maternal smoking status at antenatal booking: Non-smoker. In unadjusted models, inequality measures (SIMD quintile, parental NSSEC, urban-rural class, child ethnicity and child sex) were modelled separately in bivariate models. In adjusted models they were modelled together in the same model, along with the additional control variables (i.e. maternal age, maternal smoking status and maternal mental health hospital admissions).

**Table A3. Sensitivity analysis: odds ratios (95% confidence intervals) showing inequalities in odds of developmental concerns before, during and after the COVID-19 pandemic, Greater Glasgow and Clyde health board excluded**

|  | **Having any (i.e. at least one) developmental concern identified** | | | | | | | | | | | |
| --- | --- | --- | --- | --- | --- | --- | --- | --- | --- | --- | --- | --- |
|  | **13-15 Month Child Health Reviews** | | | | | | **27-30 Month Child Health Reviews** | | | | | |
|  | **Pre-pandemic**  **period** | | **Pandemic**  **period** | | **Post-pandemic period** | | **Pre-pandemic**  **period** | | **Pandemic**  **period** | | **Post-pandemic period** | |
|  | **Un-adjusted** | **Adjusted** | **Un-adjusted** | **Adjusted** | **Un-adjusted** | **Adjusted** | **Un-adjusted** | **Adjusted** | **Un-adjusted** | **Adjusted** | **Un-adjusted** | **Adjusted** |
| **SIMD quintile** |  |  |  |  |  |  |  |  |  |  |  |  |
| *1 (most deprived)* | 1.63  (1.47-1.80) | 1.34  (1.21-1.50) | 1.72  (1.55-1.91) | 1.43  (1.28-1.60) | 1.67  (1.55-1.80) | 1.31  (1.20-1.42) | 3.54  (3.17-3.96) | 2.25  (2.00-2.54) | 3.00  (2.71-3.32) | 1.90  (1.71-2.13) | 3.00  (2.71-3.32) | 1.84  (1.69-2.01) |
| *2 (more deprived)* | 1.37  (1.24-1.52) | 1.22  (1.09-1.36) | 1.36  (1.22-1.51) | 1.23  (1.10-1.38) | 1.41  (1.30-1.52) | 1.20  (1.11-1.30) | 2.73  (2.44-3.05) | 2.00  (1.77-2.48) | 2.28  (2.06-2.53) | 1.67  (1.49-1.86) | 2.28  (2.06-2.53) | 1.58  (1.45-1.73) |
| *3 (medium deprived)* | 1.41  (1.27-1.56) | 1.36  (1.22-1.51) | 1.33  (1.19-1.48) | 1.31  (1.18-1.47) | 1.20  (1.11-1.29) | 1.12  (1.04-1.23) | 1.92  (1.71-2.16) | 1.63  (1.44-1.84) | 1.61  (1.45-1.80) | 1.39  (1.24-1.56) | 1.61  (1.45-1.80) | 1.50  (1.37-1.64) |
| *4 (less deprived)* | 1.09  (0.98-1.21) | 1.10  (0.98-1.22) | 1.11  (1.00-1.24) | 1.14  (1.02-1.27) | 1.13  (1.05-1.22) | 1.11  (1.03-1.21) | 1.54  (1.37-1.74) | 1.47  (1.30-1.67) | 1.27  (1.14-1.42) | 1.22  (1.09-1.37) | 1.27  (1.14-1.42) | 1.26  (1.15-1.38) |
| **Parental NSSEC** |  |  |  |  |  |  |  |  |  |  |  |  |
| *Not working* | 2.27  (2.00-2.58) | 2.03  (1.76-2.34) | 2.09  (1.84-2.37) | 1.72  (1.49-1.98) | 2.37  (2.16-2.61) | 1.92  (1.73-2.13) | 4.25  (3.78-4.77) | 2.55  (2.24-2.92) | 3.71  (3.31-4.15) | 2.26  (1.99-2.57) | 3.91  (3.57-4.27) | 2.42  (2.18-2.68) |
| *Routine* | 1.52  (1.41-1.64) | 1.39  (1.27-1.52) | 1.52  (1.41-1.64) | 1.31  (1.20-1.44) | 1.56  (1.47-1.65) | 1.36  (1.27-1.45) | 2.60  (2.41-2.81) | 1.78  (1.63-1.94) | 2.53  (2.35-2.71) | 1.75  (1.61-1.90) | 2.60  (2.45-2.75) | 1.84  (1.72-1.96) |
| *Intermediate* | 1.18  (1.10-1.28) | 1.14  (1.06-1.23) | 1.09  (1.01-1.18) | 1.01  (0.94-1.10) | 1.20  (1.13-1.26) | 1.12  (1.06-1.19) | 1.75  (1.63-1.89) | 1.40  (1.30-1.52) | 1.60  (1.49-1.72) | 1.31  (1.22-1.42) | 1.72  (1.63-1.82) | 1.43  (1.35-1.52) |
| **Urban-rural class** |  |  |  |  |  |  |  |  |  |  |  |  |
| *Rural* | 0.81  (0.75-0.88) | 0.85  (0.78-0.92) | 0.78  (0.72-0.85) | 0.83  (0.77-0.91) | 0.84  (0.79-0.89) | 0.91  (0.85-0.96) | 0.72  (0.66-0.78) | 0.85  (0.78-0.93) | 0.67  (0.62-0.73) | 0.81  (0.74-0.88) | 0.74  (0.69-0.78) | 0.86  (0.81-0.92) |
| **Child ethnicity** |  |  |  |  |  |  |  |  |  |  |  |  |
| *African ethnic group* | 1.20  (0.90-1.61) | 1.10  (0.82-1.49) | 1.07  (0.76-1.51) | 1.05  (0.74-1.48) |  | 0.80  (0.62-1.02) | 1.44  (1.10-1.90) | 1.62  (1.22-2.15) | 1.26  (0.95-1.66) | 1.31  (0.98-1.74) | 1.46  (1.18-1.81) | 1.63  (1.30-2.04) |
| *Caribbean or Black ethnic group* | 1.32  (0.55-3.13) | 1.23  (0.51-2.94) | 0.66  (0.24-1.83) | 0.61  (0.22-1.71) | 0.82  (0.65-1.05) | 0.64  (0.29-1.39) | 1.13  (0.48-2.68) | 1.48  (0.61-3.59) | 0.50  (0.18-1.38) | 0.50  (0.18-1.41) | 1.25  (0.65-2.41) | 1.27  (0.64-2.51) |
| *Asian ethnic group* | 1.17  (0.99-1.39) | 1.23  (1.03-1.46) | 1.38  (1.17-1.62) | 1.48  (1.26-1.75) | 0.64  (0.29-1.40) | 1.17  (1.03-1.32) | 1.44  (1.22-1.69) | 1.74  (1.47-2.06) | 1.31  (1.12-1.53) | 1.50  (0.18-1.41) | 1.21  (1.07-1.37) | 1.47  (1.29-1.67) |
| Mixed ethnic group | 0.86  (0.70-1.05) | 0.86  (0.70-1.06) | 0.71  (0.57-0.89) | 0.72  (0.57-0.90) | 0.85  (0.73-0.99) | 0.87  (0.75-1.01) | 0.85  (0.68-1.05) | 0.94  (0.75-1.17) | 0.87  (0.72-1.05) | 1.56  (1.33-1.83) | 0.83  (0.71-0.96) | 0.91  (0.78-1.06) |
| Other ethnic group | 0.84  (0.54-1.31) | 0.71  (0.45-1.10) | 0.80  (0.50-1.29) | 0.68  (0.42-1.09) | 0.65  (0.46-0.92) | 0.57  (0.40-0.81) | 0.89  (0.55-1.44) | 0.74  (0.45-1.20) | 1.08  (0.73-1.61) | 0.94  (0.77-1.14) | 1.11  (0.82-1.49) | 0.91  (0.67-1.23) |
| **Child sex** |  |  |  |  |  |  |  |  |  |  |  |  |
| *Female* | 0.72  (0.68-0.77) | 0.72  (0.68-0.77) | 0.73  (0.69-0.78) | 0.73  (0.68-0.77) | 0.77  (0.74-0.81) | 0.77  (0.74-0.81) | 0.41  (0.39-0.44) | 0.40  (0.38-0.43) | 0.41  (0.39-0.44) | 0.40  (0.38-0.43) | 0.44  (0.42-0.45) | 0.42  (0.40-0.44) |
| ***Maternal age, years*** |  | 1.02  (1.01-1.02) |  | 1.02  (1.01-1.02) |  | 1.01  (1.00-1.01) |  | 0.99  (0.98-0.99) |  | 0.99  (0.98-0.99) |  | 0.99  (0.98-0.99) |
| **Maternal history of mental health hospital admissions** |  |  |  |  |  |  |  |  |  |  |  |  |
| *Yes* |  | 1.63  (1.37-1.94) |  | 1.70  (1.41-2.05) |  | 1.55  (1.33-1.79) |  | 1.51  (1.27-1.79) |  | 1.58  (1.34-1.86) |  | 1.42  (1.23-1.65) |
| **Maternal smoking status at antenatal booking** |  |  |  |  |  |  |  |  |  |  |  |  |
| *Ex-smoker* |  | 0.97  (0.89-1.05) |  | 1.03  (0.96-1.11) |  | 1.04  (0.98-1.10) |  | 1.14  (1.05-1.23) |  | 1.10  (1.02-1.19) |  | 1.21  (1.14-1.28) |
| *Current smoker* |  | 1.23  (1.12-1.34) |  | 1.41  (1.29-1.54) |  | 1.41  (1.32-1.51) |  | 1.45  (1.34-1.57) |  | 1.55  (1.43-1.67) |  | 1.63  (1.53-1.74) |
| *Unknown* |  | 1.43  (1.19-1.71) |  | 1.17  (0.92-1.48) |  | 0.73  (0.48-1.09) |  | 1.40  (1.12-1.75) |  | 1.39  (1.16-1.66) |  | 1.21  (0.99-1.47) |

Notes: results are from logistic regression models showing associations between each inequality measure and odds of children having any (i.e. at least one) developmental concerns. Reference categories are as follows: SIMD quintile: 5 (least deprived), Parental NSSEC: Managerial/Professional, Urban-rural class: Urban, Child ethnicity: White ethnic group, Child sex: Male, Maternal history of mental health hospital admissions: No, Maternal smoking status at antenatal booking: Non-smoker. In unadjusted models, inequality measures (SIMD quintile, parental NSSEC, urban-rural class, child ethnicity and child sex) were modelled separately in bivariate models. In adjusted models they were modelled together in the same model, along with the additional control variables (i.e. maternal age, maternal smoking status and maternal mental health hospital admissions).

**Table A4. Sensitivity analysis: Odds ratios (95% confidence intervals) showing interaction between pandemic cohort and inequality variables, Greater Glasgow and Clyde health board excluded**

|  | **Having any (i.e. at least one) developmental concern identified** | | | |
| --- | --- | --- | --- | --- |
|  | **13-15 Month Child Health Reviews** | | **27-30 Month Child Health Reviews** | |
|  | **Unadjusted** | **Adjusted** | **Unadjusted** | **Adjusted** |
| **SIMD quintile** |  |  |  |  |
| *1 (most deprived) x during pandemic* | 1.06 (0.91-1.23) | 1.09 (0.93-1.28) | 0.85 (0.73-0.99) | 0.85 (0.72-1.00) |
| *1 (most deprived) x post-pandemic* | 1.03 (0.90-1.17) | 1.01 (0.88-1.16) | 0.85 (0.74-0.97) | 0.83 (0.72-0.96) |
| *2 (more deprived) x during pandemic* | 0.99 (0.85-1.15) | 1.03 (0.88-1.20) | 0.84 (0.72-0.97) | 0.84 (0.71-0.98) |
| *2 (more deprived) x post-pandemic* | 1.02 (0.90-1.17) | 1.02 (0.89-1.16) | 0.82 (0.71-0.94) | 0.80 (0.69-0.93) |
| *3 (medium deprived) x during pandemic* | 0.94 (0.81-1.10) | 0.98 (0.84-1.14) | 0.84 (0.72-0.98) | 0.85 (0.72-1.01) |
| *3 (medium deprived) x post-pandemic* | 0.85 (0.75-0.97) | 0.85 (0.74-0.97) | 0.93 (0.80-1.07) | 0.93 (0.80-1.08) |
| *4 (less deprived) x during pandemic* | 1.02 (0.88-1.19) | 1.05 (0.89-1.22) | 0.83 (0.70-0.97) | 0.83 (0.70-0.98) |
| *4 (less deprived) x post-pandemic* | 1.04 (0.91-1.19) | 1.03 (0.89-1.18) | 0.87 (0.75-1.01) | 0.86 (0.74-1.00) |
| **Parental NSSEC** |  |  |  |  |
| *Not working x during pandemic* | 0.92 (0.77-1.10) | 0.90 (0.75-1.09) | 0.87 (0.74-1.03) | 0.90 (0.76-1.07) |
| *Not working x post-pandemic* | 1.05 (0.89-1.22) | 1.03 (0.88-1.22) | 0.92 (0.79-1.07) | 0.98 (0.84-1.14) |
| *Routine x during pandemic* | 1.00 (0.90-1.22) | 0.98 (0.87-1.11) | 0.97 (0.88-1.080 | 0.99 (0.88-1.11) |
| *Routine x post-pandemic* | 1.03 (0.93-1.14) | 1.04 (0.93-1.15) | 1.00 (0.91-1.10) | 1.06 (0.95-1.17) |
| *Intermediate x during pandemic* | 0.92 (0.83-1.02) | 0.91 (0.81-1.01) | 0.91 (0.82-1.01) | 0.94 (0.84-1.04) |
| *Intermediate x post-pandemic* | 1.01 (0.92-1.11) | 1.02 (0.92-1.12) | 0.98 (0.90-1.08) | 1.03 (0.94-1.14) |
| **Urban-rural class** |  |  |  |  |
| *Rural x during pandemic* | 0.96 (0.86-1.08) | 0.98 (0.86-1.10) | 0.93 (0.83-1.04) | 0.95 (0.84-1.07) |
| *Rural x post-pandemic* | 1.04 (0.93-1.15) | 1.06 (0.95-1.18) | 1.02 (0.92-1.13) | 1.01 (0.90-1.13) |
| **Child ethnicity** |  |  |  |  |
| *African ethnic group x during pandemic* | 0.89 (0.57-1.40) | 0.91 (0.58-1.43) | 0.87 (0.59-1.29) | 0.81 (0.54-1.21) |
| *African ethnic group x post-pandemic* | 0.68 (0.47-1.00) | 0.68 (0.46-1.00) | 1.01 (0.71-1.43) | 0.98 (0.68-1.41) |
| *Caribbean or Black ethnic group x during pandemic* | 0.50 (0.13-1.90) | 0.48 (0.12-1.84) | 0.44 (0.12-1.67) | 0.34 (0.09-1.32) |
| *Caribbean or Black ethnic group x post-pandemic* | 0.49 (0.15-1.56) | 0.49 (0.15-1.58) | 1.10 (0.37-3.25) | 0.84 (0.28-2.58) |
| *Asian ethnic group x during pandemic* | 1.17 (0.93-1.49) | 1.17 (0.92-1.48) | 0.91 (0.73-1.14) | 0.89 (0.71-1.13) |
| *Asian ethnic group x post-pandemic* | 0.94 (0.76-1.16) | 0.92 (0.74-1.13) | 0.84 (0.69-1.03) | 0.82 (0.66-1.01) |
| Mixed ethnic group *x during pandemic* | 0.83 (0.61-1.13) | 0.83 (0.61-1.13) | 1.02 (0.77-1.36) | 1.00 (0.75-1.34) |
| Mixed ethnic group *x post-pandemic* | 0.99 (0.77-1.28) | 1.00 (0.77-1.28) | 0.98 (0.75-1.27) | 0.96 (0.74-1.26) |
| Other ethnic group *x during pandemic* | 0.95 (0.50-1.82) | 0.91 (0.47-1.74) | 1.21 (0.65-2.26) | 1.33 (0.71-2.51) |
| Other ethnic group *x post-pandemic* | 0.77 (0.44-1.35) | 0.76 (0.43-1.33) | 1.24 (0.71-2.17) | 1.19 (0.67-2.11) |
| **Child sex** |  |  |  |  |
| *Female x during pandemic* | 1.01 (0.92-1.10) | 1.01 (0.92-1.10) | 1.00 (0.92-1.09) | 1.00 (0.91-1.09) |
| *Female x post-pandemic* | 1.07 (0.99-1.16) | 1.07 (0.99-1.16) | 1.06 (0.98-1.15) | 1.05 (0.97-1.14) |

Notes: results are from logistic regression models showing showing interaction between pandemic cohort and inequality variables. Reference categories are as follows: Pandemic cohort: Pre-pandemic, SIMD quintile: 5 (least deprived), Parental NSSEC: Managerial/Professional, Urban-rural class: Urban, Child ethnicity: White ethnic group, Child sex: Male, Maternal history of mental health hospital admissions: No, Maternal smoking status at antenatal booking: Non-smoker. In unadjusted models, inequality measures (SIMD quintile, parental NSSEC, urban-rural class, child ethnicity and child sex) were modelled separately. In adjusted models they were modelled together in the same model, which additionally adjusted for the additional control variables (i.e. maternal age, maternal smoking status and maternal mental health hospital admissions).

**Table A5. Sensitivity analysis: odds ratios (95% confidence intervals) showing inequalities in odds of developmental concerns before, during and after the COVID-19 pandemic, models including gestational age at birth as additional control variable**

|  | **Having any (i.e. at least one) developmental concern identified** | | | | | |
| --- | --- | --- | --- | --- | --- | --- |
|  | **13-15 Month Child Health Reviews** | | | **27-30 Month Child Health Reviews** | | |
|  | **Pre-pandemic**  **period** | **Pandemic**  **period** | **Post-pandemic period** | **Pre-pandemic**  **period** | **Pandemic**  **period** | **Post-pandemic period** |
|  | **Adjusted (including gestational age at birth)** | **Adjusted (including gestational age at birth)** | **Adjusted (including gestational age at birth)** | **Adjusted (including gestational age at birth)** | **Adjusted (including gestational age at birth)** | **Adjusted (including gestational age at birth)** |
| **SIMD quintile** |  |  |  |  |  |  |
| *1 (most deprived)* | 1.12 (1.01-1.25) | 1.13 (1.02-1.26) | 1.09 (1.01-1.18) | 2.22 (2.00-2.47) | 1.97 (1.79-2.16) | 1.93 (1.79-2.08) |
| *2 (more deprived)* | 1.18 (1.06-1.31) | 1.20 (1.08-1.33) | 1.18 (1.09-1.27) | 1.94 (1.74-2.16) | 1.63 (1.49-1.79) | 1.59 (1.47-1.71) |
| *3 (medium deprived)* | 1.33 (1.20-1.49) | 1.31 (1.18-1.46) | 1.15 (1.06-1.24) | 1.59 (1.42-1.78) | 1.35 (1.22-1.49) | 1.46 (1.35-1.57) |
| *4 (less deprived)* | 1.09 (0.97-1.21) | 1.14 (1.03-1.27) | 1.15 (1.07-1.24) | 1.43 (1.28-1.60) | 1.18 (1.07-1.31) | 1.23 (1.14-1.32) |
| **Parental NSSEC** |  |  |  |  |  |  |
| *Not working* | 1.85 (1.61-2.12) | 1.68 (1.48-1.91) | 1.82 (1.65-2.00) | 2.29 (2.05-2.57) | 1.96 (1.76-2.18) | 2.15 (1.97-2.33) |
| *Routine* | 1.36 (1.24-1.48) | 1.34 (1.23-1.46) | 1.40 (1.31-1.49) | 1.68 (1.55-1.81) | 1.67 (1.55-1.79) | 1.70 (1.60-1.79) |
| *Intermediate* | 1.12 (1.03-1.21) | 1.03 (0.96-1.12) | 1.14 (1.08-1.21) | 1.34 (1.25-1.43) | 1.32 (1.23-1.40) | 1.39 (1.32-1.46) |
| **Urban-rural class** |  |  |  |  |  |  |
| *Rural* | 0.95 (0.87-1.04) | 0.96 (0.88-1.04) | 1.04 (0.97-1.10) | 0.82 (0.75-0.89) | 0.78 (0.72-0.84) | 0.81 (0.76-0.86) |
| **Child ethnicity** |  |  |  |  |  |  |
| *African ethnic group* | 0.83 (0.63-1.09) | 0.72 (0.54-0.95) | 0.64 (0.53-0.78) | 1.57 (1.27-1.95) | 1.32 (1.09-1.60) | 1.69 (1.47-1.94) |
| *Caribbean or Black ethnic group* | 0.98 (0.42-2.31) | 0.74 (0.32-1.71) | 0.49 (0.53-0.78) | 1.42 (0.71-2.85) | 1.00 (0.52-1.93) | 1.48 (0.89-2.46) |
| *Asian ethnic group* | 0.92 (0.78-1.10) | 0.98 (0.85-1.14) | 0.49 (0.23-1.06) | 1.68 (1.47-1.91) | 1.50 (1.33-1.68) | 1.51 (1.38-1.65) |
| Mixed ethnic group | 0.84 (0.69-1.02) | 0.68 (0.55-0.84) | 0.85 (0.75-0.98) | 1.03 (0.86-1.24) | 1.04 (0.89-1.22) | 0.96 (0.85-1.09) |
| Other ethnic group | 0.50 (0.33-0.76) | 0.49 (0.33-0.72) | 0.46 (0.35-0.59) | 1.06 (0.80-1.41) | 1.34 (1.06-1.69) | 1.00 (0.84-1.20) |
| **Child sex** |  |  |  |  |  |  |
| *Female* | 0.73 (0.69-0.78) | 0.72 (0.68-0.77) | 0.78 (0.74-0.81) | 0.41 (0.38-0.43) | 0.41 (0.39-0.43) | 0.43 (0.42-0.45) |
| **Maternal age, years** | 1.01 (1.00-1.02) | 1.01 (1.00-1.01) | 1.00 (1.00-1.01) | 0.99 (0.98-0.99) | 0.99 (0.98-0.99) | 0.99 (0.98-0.99) |
| **Maternal history of mental health hospital admissions** |  |  |  |  |  |  |
| *Yes* | 1.55 (1.31-1.85) | 1.55 (1.30-1.85) | 1.48 (1.29-1.70) | 1.44 (1.23-1.68) | 1.46 (1.26-1.68) | 1.29 (1.13-47) |
| **Maternal smoking status at antenatal booking** |  |  |  |  |  |  |
| *Ex-smoker* | 1.07 (0.99-1.16) | 1.11 (1.03-1.19) | 1.12 (1.06-1.19) | 1.15 (1.07-1.24) | 1.10 (1.03-1.17) | 1.19 (1.13-1.25) |
| *Current smoker* | 1.27 (1.16-1.38) | 1.44 (1.33-1.57) | 1.47 (1.38-1.57) | 1.40 (1.30-1.50) | 1.51 (1.41-1.61) | 1.55 (1.47-1.64) |
| *Unknown* | 1.37 (1.15-1.64) | 1.25 (0.98-1.59) | 0.77 (0.52-1.13) | 1.29 (1.06-1.58) | 1.22 (1.05-1.43) | 1.07 (0.88-1.30) |
| **Gestational age at birth, weeks** | 0.89 (0.88-0.91) | 0.89 (0.88-0.90) | 0.90 (0.89-0.91) | 0.91 (0.90-0.92) | 0.91 (0.90-0.92) | 0.91 (0.90-0.92) |

Notes: results are from logistic regression models showing associations between each inequality measure and odds of children having any (i.e. at least one) developmental concerns. Reference categories are as follows: SIMD quintile: 5 (least deprived), Parental NSSEC: Managerial/Professional, Urban-rural class: Urban, Child ethnicity: White ethnic group, Child sex: Male, Maternal history of mental health hospital admissions: No, Maternal smoking status at antenatal booking: Non-smoker.

**Table A6. Sensitivity analysis: Odds ratios (95% confidence intervals) showing interaction between pandemic cohort and inequality variables models including gestational age at birth as additional control variable**

|  | **Having any (i.e. at least one) developmental concern identified** | |
| --- | --- | --- |
|  | **13-15 Month Child Health Reviews** | **27-30 Month Child Health Reviews** |
|  | **Adjusted (including gestational age at birth)** | **Adjusted (including gestational age at birth)** |
| **SIMD quintile** |  |  |
| *1 (most deprived) x during pandemic* | 1.04 (0.89-1.21) | 0.89 (0.77-1.02) |
| *1 (most deprived) x post-pandemic* | 1.01 (0.89-1.15) | 0.88 (0.78-1.00) |
| *2 (more deprived) x during pandemic* | 1.04 (0.89-1.21) | 0.84 (0.73-0.97) |
| *2 (more deprived) x post-pandemic* | 1.03 (0.91-1.18) | 0.83 (0.73-0.94) |
| *3 (medium deprived) x during pandemic* | 0.99 (0.85-1.15) | 0.85 (0.73-0.98) |
| *3 (medium deprived) x post-pandemic* | 0.88 (0.77-1.00) | 0.92 (0.81-1.06) |
| *4 (less deprived) x during pandemic* | 1.06 (0.91-1.23) | 0.83 (0.71-0.96) |
| *4 (less deprived) x post-pandemic* | 1.07 (0.94-1.22) | 0.86 (0.75-0.98) |
| **Parental NSSEC** |  |  |
| *Not working x during pandemic* | 0.97 (0.82-1.16) | 0.86 (0.74-1.00) |
| *Not working x post-pandemic* | 1.08 (0.92-1.26) | 0.96 (0.84-1.10) |
| *Routine x during pandemic* | 1.03 (0.92-1.16) | 1.00 (0.90-1.10) |
| *Routine x post-pandemic* | 1.10 (0.99-1.21) | 1.04 (0.95-1.14) |
| *Intermediate x during pandemic* | 0.95 (0.86-1.05) | 0.98 (0.90-1.08) |
| *Intermediate x post-pandemic* | 1.06 (0.97-1.16) | 1.05 (0.97-1.15) |
| **Urban-rural class** |  |  |
| *Rural x during pandemic* | 1.00 (0.89-1.14) | 0.95 (0.85-1.07) |
| *Rural x post-pandemic* | 1.08 (0.97-1.20) | 0.99 (0.89-1.10) |
| **Child ethnicity** |  |  |
| *African ethnic group x during pandemic* | 0.83 (0.56-1.22) | 0.84 (0.63-1.12) |
| *African ethnic group x post-pandemic* | 0.72 (0.52-1.01) | 1.04 (0.81-1.34) |
| *Caribbean or Black ethnic group x during pandemic* | 0.72 (0.22-2.39) | 0.70 (0.27-1.82) |
| *Caribbean or Black ethnic group x post-pandemic* | 0.47 (0.15-1.48) | 1.02 (0.43-2.40) |
| *Asian ethnic group x during pandemic* | 1.03 (0.83-1.28) | 0.89 (0.75-1.06) |
| *Asian ethnic group x post-pandemic* | 1.01 (0.83-1.23) | 0.88 (0.75-1.06) |
| Mixed ethnic group *x during pandemic* | 0.80 (0.60-1.07) | 1.01 (0.79-1.28) |
| Mixed ethnic group *x post-pandemic* | 1.00 (0.78-1.26) | 0.92 (0.74-1.16) |
| Other ethnic group *x during pandemic* | 0.93 (0.53-1.62) | 1.24 (0.86-1.79) |
| Other ethnic group *x post-pandemic* | 0.86 (0.53-1.39) | 0.91 (0.65-1.27) |
| **Child sex** |  |  |
| *Female x during pandemic* | 0.98 (0.90-1.07) | 1.00 (0.93-1.08) |
| *Female x post-pandemic* | 1.06 (0.98-1.14) | 1.07 (1.00-1.15) |

Notes: results are from logistic regression models showing showing interaction between pandemic cohort and inequality variables. Reference categories are as follows: Pandemic cohort: Pre-pandemic, SIMD quintile: 5 (least deprived), Parental NSSEC: Managerial/Professional, Urban-rural class: Urban, Child ethnicity: White ethnic group, Child sex: Male, Maternal history of mental health hospital admissions: No, Maternal smoking status at antenatal booking: Non-smoker. Models adjust for maternal age, maternal smoking status, maternal mental health hospital admissions and gestational age at birth.
